# Distinct baseline immune characteristics associated with responses to conjugated and unconjugated pneumococcal polysaccharide vaccines in older adults

**DOI:** 10.1101/2023.04.16.23288531

**Authors:** Sathyabaarathi Ravichandran, Fernando Erra-Diaz, Onur E. Karakaslar, Radu Marches, Lisa Kenyon-Pesce, Robert Rossi, Damien Chaussabel, Virginia Pascual, Karolina Palucka, Silke Paust, Moon H. Nahm, George A. Kuchel, Jacques Banchereau, Duygu Ucar

## Abstract

Pneumococcal infections cause serious illness and death among older adults. A capsular polysaccharide vaccine PPSV23 (Pneumovax®) and a conjugated polysaccharide vaccine PCV13 (Prevnar®) are used to prevent these infections, yet underlying responses, and baseline predictors remain unknown. We recruited and vaccinated 39 older adults (>60 years) with PPSV23 or PCV13. Both vaccines induced strong antibody responses at day 28 and similar plasmablast transcriptional signatures at day 10, however, their baseline predictors were distinct. Analyses of baseline flow cytometry and RNA-seq data (bulk and single cell) revealed a novel baseline phenotype that is specifically associated with weaker PCV13 responses, characterized by i) increased expression of cytotoxicity-associated genes and increased CD16^+^ NK frequency; ii) increased T_h_17 and decreased T_h_1 cell frequency. Men were more likely to display this cytotoxic phenotype and mounted weaker responses to PCV13 than women. Baseline expression levels of a distinct gene set was predictive of PPSV23 responses. This first precision vaccinology study for pneumococcal vaccine responses of older adults uncovered novel and distinct baseline predictors that might transform vaccination strategies and initiate novel interventions.

## INTRODUCTION

*Streptococcus pneumoniae* (Pneumococcus) infections can lead to life-threatening diseases including meningitis, pneumonia, and sepsis, and are estimated to be responsible for 445,000 hospitalizations and 22,000 deaths annually in the US^(1, 2)^. Infants and older adults have the highest rates of invasive pneumococcal disease (IPD). IPD has an overall mortality rate of ∼10%^(3)^, with case-fatality rates further increasing with age for reasons that are not well understood. Two types of pneumococcal vaccines which elicit neutralizing antibodies to *Streptococcus pneumoniae* are FDA-approved for clinical use in the US: the capsular polysaccharide vaccine PPSV23 (Pneumovax®) and the protein-polysaccharide conjugate vaccines (e.g., Prevnar® - PCV13). PPSV23 contains capsular polysaccharide antigens for 23 serotypes (1, 2, 3, 4, 5, 6B, 7F, 8, 9N, 9V, 10A, 11A, 12F, 14, 15B, 17F, 18C, 19A, 19F, 20, 22F, 23F, and 33F) and induces T-cell-independent antibody responses^(4)^. To enhance immunogenicity, conjugated vaccines have been developed (e.g., PCV13). PCV13 is composed of capsular polysaccharides of 13 serotypes of *Streptococcus pneumoniae* (1, 3, 4, 5, 6A, 6B, 7F, 9V, 14, 19A, 19F, 18C, and 23F) conjugated to a nontoxic variant of diphtheria toxin CRM197 that induces T cell responses, thus also providing T cell help in vaccine-induced antibody production^(5, 6)^.

Conjugated pneumococcal vaccines such as PCV13 are effective among children and young adults^(7)^. They are also effective in preventing invasive disease among older adults^(8)^, however, their protection declines with age for reasons that are not well-defined^(9–11)^. In 2021, other conjugated vaccines were approved for use: PCV15 (Vaxneuvance®) and PCV20 (Prevnar® 20), in hopes of further improving immunogenicity and simplifying pneumococcal immunizations^(12)^. In the US, the current recommendation for adults aged ≥ 65 years is to give a conjugated vaccine (PCV15/PCV20) followed by PPSV23 at least one year later. In addition, patients with a high-risk condition (i.e., immuno-compromised, chronic medical conditions) are recommended to receive one dose of PCV20 or a combination of PCV15 and PPSV23. Recommendations for pneumococcal vaccines have been complex and changing frequently, which likely contributes to suboptimal uptake of the vaccines. Hence, there is an unmet clinical need to identify baseline predictors for vaccine responsiveness for a more targeted and precise match between a patient and a specific pneumococcal vaccine^(13, 14)^.

Peripheral blood leukocyte gene expression studies by us and others of pneumococcal vaccination identified transcriptional signatures of vaccine responses^(15, 16)^ and showed that healthy adults (18-64 years old) respond to PPSV23 differently than to the seasonal influenza vaccine^(16)^. However, these studies did not include conjugated pneumococcal vaccine and excluded older adults. To address these gaps in knowledge, we recruited 39 healthy pneumococcal vaccine-naïve adults ≥ 60 years old and vaccinated them with either PCV13 (n=19) or with PPSV23 (n=20) during the 2017 and 2018 seasons. We collected blood from this cohort longitudinally and studied their baseline immune phenotypes and immune responses upon vaccination using multiple assays: opsonophagocytic antibody assay (OPA) to quantify functional antibody responses, flow cytometry, as well as PBMC bulk and single cell RNA-seq. Our systems vaccinology approach uncovered a baseline activated immune phenotype that is specifically associated with weaker PCV13 responses. Donors who had this baseline phenotype had a higher T_h_17 frequency and a lower T_h_1 frequency as well as increased expression of cytotoxicity-associated genes and increased frequency of CD16^+^ NK cells; this phenotype was specifically associated with the conjugated PCV13 vaccine responses. In contrast, we uncovered a distinct baseline gene expression module associated with reduced PPSV23 responses, which included 134 genes associated with cell cycle and transcriptional regulation.

## RESULTS

### Serological responses of PCV13 and PPSV23 vaccines in older adults

We measured and compared the immune responses of healthy older adults to two pneumococcal vaccines: the 13-valent T-dependent conjugate PCV13 and the 23-valent polysaccharide T-independent PPSV23. 39 older adults (≥ 60 years of age) who had not been previously vaccinated with any pneumococcal vaccine were recruited and randomized to receive a single dose of PCV13 (10 men, 9 women) or PPSV23 (10 men, 10 women) during the 2017 and 2018 seasons. Volunteers were vaccinated from May to early fall to avoid overlap with potentially confounding peak periods of influenza vaccination and infection. None of the participants reported a history of a previous pneumonia. Individuals receiving PCV13 or PPSV23 were comparable in terms of age, Body Mass Index (BMI), Cytomegalovirus (CMV) seropositivity and Frailty Index. The average age in the cohort was 68 years (**Supplementary Table 1a, 1b,** **Fig. 1a**). Furthermore, men and women in each cohort were comparable in terms of age, BMI, CMV seropositivity and Frailty Index. (**Supplementary Table 1b**).

**Figure 1:**
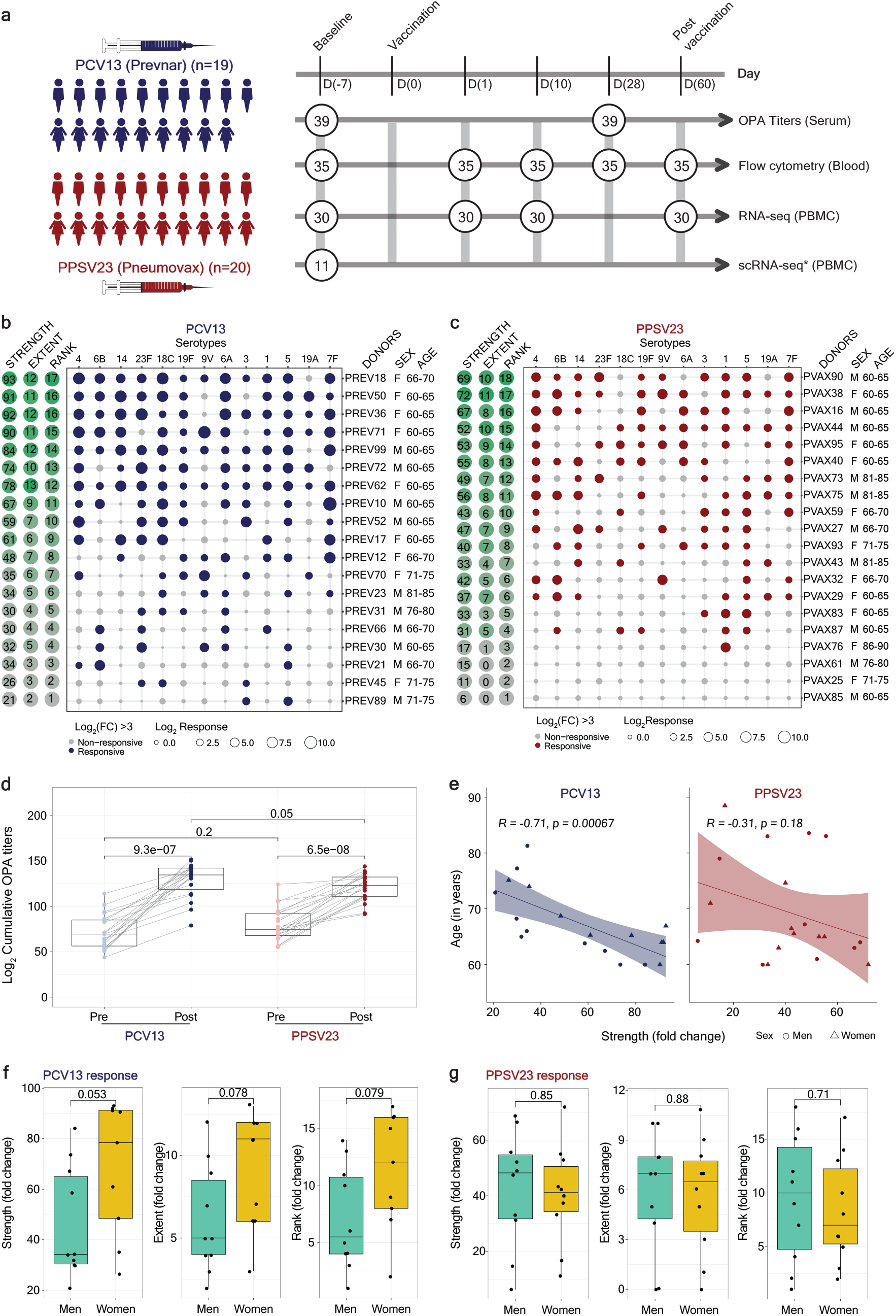
Functional antibody response to PCV13 and PPSV23 in older adults. **a**) Schematic representation of the study design. 9 women and 10 men received PCV13 vaccine, and 10 women and 10 men received PPSV23. OPA titers for the 13 serotypes were assessed from serum samples obtained 7 days prior to vaccination (baseline), and 28 days post-vaccination, for both vaccines. Anticoagulated blood samples were used for flow cytometric analysis of whole blood cell populations. PBMCs (Peripheral Blood Mononuclear Cells) were isolated for bulk RNA-seq. Pre-vaccination PBMCs from 4 women and 7 men who received PCV13 were isolated for scRNA-sequencing. Numbers in circles represent total number of samples processed for the indicated assay at the indicated time. **b**) Bubble plot of fold change (FC) in antibody titers for individual serotypes in response to PCV13. **c**) Bubble plot of FC in antibody titers for individual serotypes in response to PPSV23. Size of the dots represent the FC value, and color indicates significant response (Log_2_ FC is > 3), with blue for PCV13 and red for PPSV23. Donors are ordered from top to bottom according to the vaccine response Rank. On the left side, the Strength (Log_2_ Sum FC), the Extent of the response (number of serotypes out of 13 to which an individual mounted significant response) and Rank are presented. **d**) Pre-vaccination and post-vaccination cumulative OPA titers (expressed as sum Log_2_). **e**) Correlation analysis between the cumulative FC (Sum Log_2_ FC) and age (in years). Sex-specific differences in the Strength, Extent and Rank in donors who received PCV13 (**f**) and PPSV23 (**g**). The Wilcoxon matched-pairs signed-rank test was used in (d) to compare Pre- and Post-vaccination titers in PCV13 and PPSV23. Wilcoxon Rank sum test was used to compare the Strength, Extent and Rank between men and women in PCV13 and PPSV23. The Pearson correlation metric was used to perform correlation analysis between Strength and age (e).

Peripheral blood samples were collected 7 days before vaccination (baseline), as well as 1, 10, 28, and 60 days after vaccination. We generated longitudinal flow cytometry and RNA-seq data for an in-depth study of the immune responses to these two vaccines (**Fig. 1a**). To evaluate vaccine-specific antibody responses and functionality, each donor’s serum opsonophagocytic activity (OPA) was quantified against 13 serotypes (12 shared serotypes and serotype 6A from PCV13) at baseline (day -7) and 28 days post-vaccination (day 28) **(Supplementary Table 2)**^(17, 18)^. OPA measures the ability of antibodies to effectively opsonize bacteria leading to their killing and is ideal for quantifying responses to pneumococcal vaccines since it mimics the *in vivo* mechanisms of antibody protection^(19)^. Typically, a response to a serotype is considered positive and significant if vaccination induces at least eight-fold increase in baseline OPA titer levels^(20)^. For multivariate analyses of responses to the different serotypes, we developed three measures: i) **Strength** of the response - the sum of fold changes (FC) between baseline and day 28 for all serotypes; ii) **Extent** - the number of serotypes (out of 13) to which the donor elicits significant (*i.e.*, >= 8-fold increase^(21)^) responses; and iii) **Rank** – the rank of an individual’s vaccine responsiveness in the cohort based on aggregate responses in OPA titers for all serotypes, where higher ranks represent the most responsive donors (**Fig. 1b** for PCV13 and **Fig. 1c** for PPSV23, **Fig. s1a,b** for other scores, **Supplementary Table 2d, 2h**). This multivariate ranking strategy enabled combining responses for all serotypes into a single responsiveness measure (i.e., rank), which correlated significantly with the vaccine responsiveness computed using the maximum residual after baseline adjustment (maxRBA) approach^(22)^ for both PCV13 (r=0.83, p=1.1e-15) and PPSV23 (r=0.8, p=2.4e-05) (**Fig. s2a**). As expected, strength and extent of responsiveness were also highly correlated with the rank, while capturing different aspects of vaccine responses (**Fig. s2a**).

Upon vaccination, OPA titer levels significantly increased for both vaccines cumulatively (**Fig. 1d**) and for each serotype **(Fig. s2b**). Significantly stronger PCV13 responses were detected only for the shared serotype 4 and as expected for the PCV13-specific serotype 6A (**Fig. s1b**). Although PPSV23 does not contain serotype 6A, there were increased responses to this serotype also in the PPSV23 cohort potentially due to the cross-reactivity between serotypes 6A and 6B^(20)^. Baseline OPA titers were comparable between the two cohorts (p=0.2), however, individuals vaccinated with PCV13 had slightly stronger responses when compared to individuals who received PPSV23 (**Fig. 1d**, p=0.05, Wilcoxon rank-sum test) even after excluding the serotype 6A (p=0.089, **Fig. s1c**). Although all donors elicited immune responses to these vaccines, level of responsiveness varied from one individual to another. In PCV13, top responder PREV18 (F, 66 - 70 years old) responded significantly to all serotypes except for 19A, whereas the bottom responder PREV89 (M, 71 - 75 years old) elicited significant responses to only two serotypes (3 and 5) (**Fig. 1b**). In PPSV23, top responder PVAX90 (M, 61 - 65 years old) responded to 10 serotypes out of the 12 that are in this vaccine, while the bottom responder PVAX85 (M, 61 - 65 years old) did not respond significantly to any of the tested serotypes (**Fig. 1c**). Our ranking strategy captured this heterogeneity at the donor level while providing a systematic way to uncover what separates responders from non-responders.

Frailty index and BMI were not significantly associated with this variation (**Fig. s2d**) (**Supplementary Table 1a, 1b**). Age of the donors was negatively associated with both vaccine responsiveness and the strength of these responses: Pearson R = -0.71, p= 0.00067 for PCV13, Pearson R: -0.31, p= 0.18 for PPSV23 (**Fig. 1e**, **Fig. s2e** for rank). Biological sex was also a factor contributing to the PCV13 vaccine responses, whereby women developed stronger responses to this vaccine compared to men (p=0.053 for the strength of response, p=0.079 for rank, **Fig. 1f**). Sex was not a contributing factor for PPSV23 responsiveness (p=0.85 for strength and p=0.71 for rank, **Fig. 1g**). Together, these data showed that both pneumococcal vaccines induced strong antibody responses; yet, at the individual level, the degree of responsiveness was variable. For PCV13, donors’ age and sex contributed to this variation, with younger donors and women mounting stronger responses to this vaccine. Interestingly, neither age nor sex was significantly associated with PPSV23 responsiveness.

### PCV13 and PPSV23 induce plasmablast transcriptional response 10 days after vaccination

To compare gene expression profiles of circulating immune cells before and after vaccination, we generated longitudinal bulk RNA-seq data (baseline, day 1, day 10, day 60) from peripheral blood mononuclear cells (PBMCs) of 31 donors from our cohort (15 vaccinated with PCV13, 16 with PPSV23). These samples were selected to include the best and worst responders for each vaccine type, while also balancing female and male samples. Samples from 30 donors that passed quality control were used in differential analyses. We compared gene expression profiles after vaccination (day 1, day 10, day 60) with the baseline profiles (day -7) (**Supplementary Table 3**). For both vaccines, we did not detect any statistically significant changes in gene expression levels for days 1 or 60 when compared to the baseline. However, at day 10, we detected 50 and 41 genes that were differentially expressed with respect to baseline (FDR < 0.05) respectively for PCV13 and PPSV23 (**Fig. s3a, Supplementary Table 3b, 3e**). Upregulated genes upon vaccination included immunoglobulin heavy chain genes *IGHG2*, *IGHA2* and plasmablast-associated genes including *JCHAIN* and *MZB1*^(23)^ (**Fig. 2a**, **Supplementary Table 3b, 3e**). Further inspection of genes associated with plasmablasts revealed that their activation peaked at day 10 and returned to baseline levels by day 60 (**Fig. 2b**).

**Figure 2:**
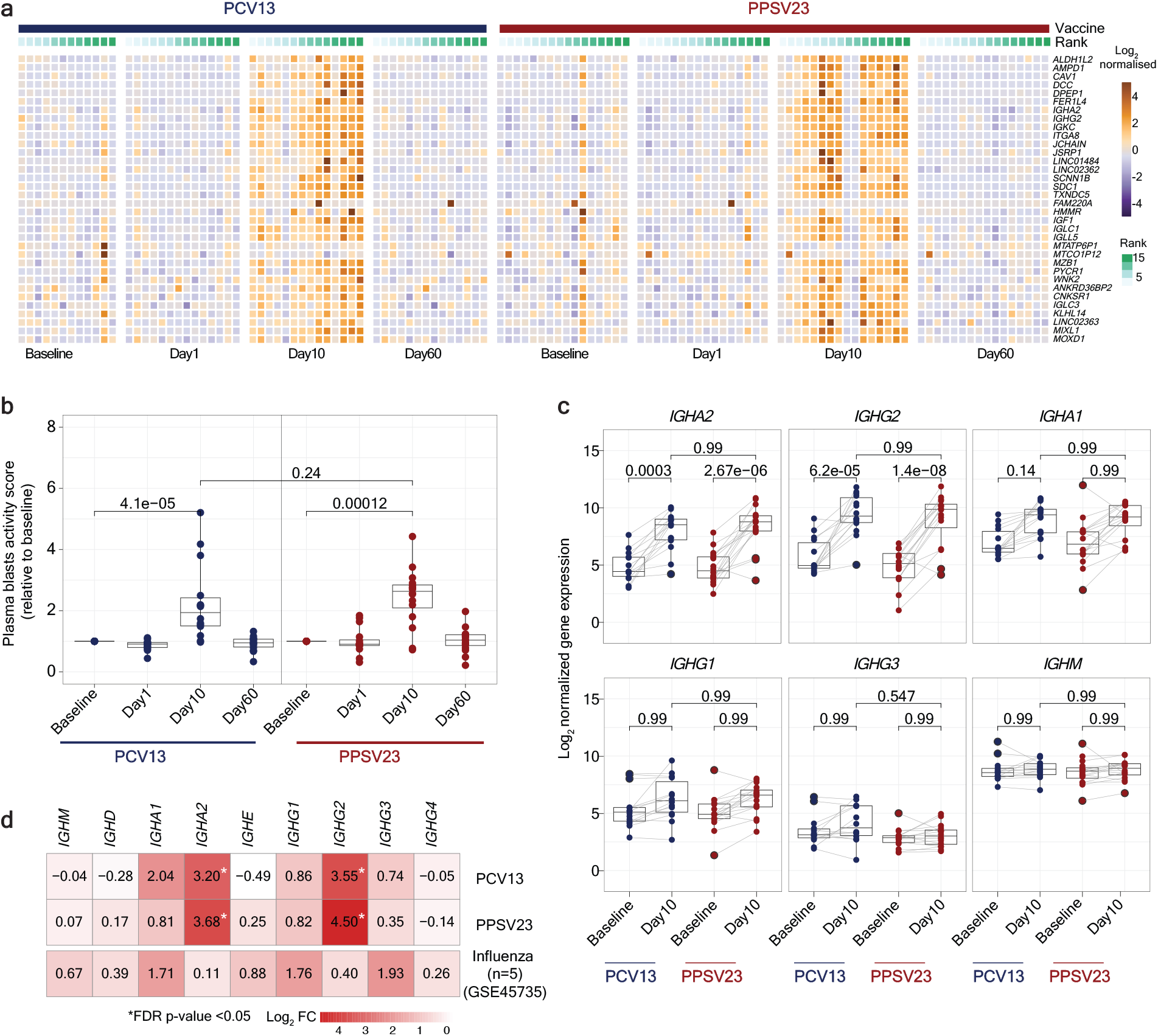
Plasmablasts response elicited upon vaccination at Day 10 in PBMCs. **a**) Heatmap of differentially expressed genes between Day 10 and baseline, using normalized gene expression values. **b**) Boxplot of plasmablasts activity scores at baseline, Day 1, Day 10, and Day 60, calculated using a published gene set (M4.11), and scaled with reference to baseline. **c**) Boxplots of normalized expression of genes coding for the constant region of immunoglobulin heavy chain structure. **d**) Heatmap showing differential expression of genes coding for the constant region of immunoglobulin heavy chain structure at Day 10 in response to PCV13, PPSV23, and Day 7 in response to Fluzone (GSE45735 - Influenza vaccine). Genes with a fold change >1.5-fold difference and FDR corrected p-value <0.05 are marked with a star. The Wilcoxon Rank sum test was used to compare plasma cell activity scores between baseline and Day 1, Day 10, and Day 60 (b). The Wilcoxon matched-pairs signed-rank test was used to compare the expression of immunoglobulin genes at baseline and Day 10 for PCV13 and PPSV23. FDR corrected p-value is shown (c).

To cumulatively quantify the plasmablast transcriptomic signature, we computed a plasmablast activity score using plasma-cell specific genesets^(16)^ (n=33 genes), which confirmed a significant (p= 4.1e-05 in PCV13 and p=0.00012 in PPSV23) and comparable increase in the expression of plasmablast-associated genes at day 10 for both vaccines and their return to baseline levels at day 60 (**Fig. 2b**). These findings are consistent with our previous study, where we observed upregulation of gene-sets associated with plasmablast response and an increase in the absolute number of plasmablasts on day 7 in young adults in response to PPSV23^(16)^. As expected^(24)^, *IGHG2,* which encodes for the IgG2 isotype, was among the most differentially expressed molecules and was similarly upregulated by both vaccines (**Fig. 2c**). We detected significant upregulation of *IGHA2* gene that encode for IgA2 isotype; this gene was similarly upregulated by both vaccines (**Fig. 2c, Fig. s3b**), in alignment with the previously reported serology data 30 days post vaccination^(25)^.

Pneumococcal vaccines upregulated distinct immunoglobulin genes compared to the seasonal influenza vaccine in agreement with the differential induction of IgG isotypes by different vaccines and infections^(26)^. Re-analysis of previously published PBMC RNA-seq data^(27)^ (n=5) showed that influenza vaccine mostly upregulated *IGHG1* and *IGHG3* genes at day 7, whereas pneumococcal vaccines upregulated *IGHA2* and *IGHG2* (**Fig. 2d, Fig. s3c**). Transcriptional activation of plasmablast-associated genes significantly correlated with PCV13 responsiveness (R=0.57, p=0.034), mainly driven by *IGHG2* expression (**Fig. s3d, f**). However, we did not detect this association for PPSV23 responses (**Fig. s3e, f**), which might be due to the quantification of responses for only 13 out of 23 serotypes included in this vaccine. In sum, transcriptional profiling before and after vaccination revealed that both pneumococcal vaccines induced significant and similar plasmablast responses 10 days post-vaccination, including the increased transcription of genes encoding for IgG2 and IgA2 antibody isotypes, contrasting with those (IgG1, IgG3, and IgA1) induced by the influenza vaccine.

### Baseline frequency of T_h_1 and T_h_17 cells are significantly associated with PCV13 vaccine responsiveness

We performed a longitudinal flow cytometry analysis of different cell populations in freshly isolated PBMCs collected from 35 donors at baseline (day -7), and post-vaccination days 1, 10, 28 and 60 to study B cell subsets (naive, memory, and plasmablast), CD4^+^ T cell naïve and memory subsets (T_h_1, T_h_2, T_h_17, T_h_10, Tfh, Tfh1, Tfh2, and Tfh17), as well as dendritic cell subsets (cDC2, cDC1, and pDC) (**Supplementary Table S4a,** **Fig 1a, Fig. s4a**). Despite the robust plasmablast transcriptional signature, their absolute numbers were only marginally increased at day 10 for both vaccines when compared to baseline (**Fig. 3a** top). We did not detect any significant changes in other B cell subsets upon vaccination or between the two cohorts (**Fig. s4b**). Among the studied T cell subsets, only ICOS^+^ Tfh cells significantly increased at day 10 for both vaccines in terms of cell numbers when compared to baseline (**Fig. 3a** bottom, **Fig. s4b**). There were no significant changes in cell numbers for other T cell or DC subsets upon vaccination (**Fig. 3b**). Furthermore, cell numbers were similar between the two cohorts for all of these profiled cell types both at baseline and upon vaccination (**Fig. s4b**). In sum, among all tested cell types, the most significant changes upon vaccination included an increase in plasmablasts and ICOS^+^ Tfh cells at day 10, which was comparable between the two vaccines. However, unlike our previous observations for influenza vaccine responses^(28)^, changes in these cell types did not correlate with level of vaccine responsiveness (**Fig. s4c**).

**Figure 3:**
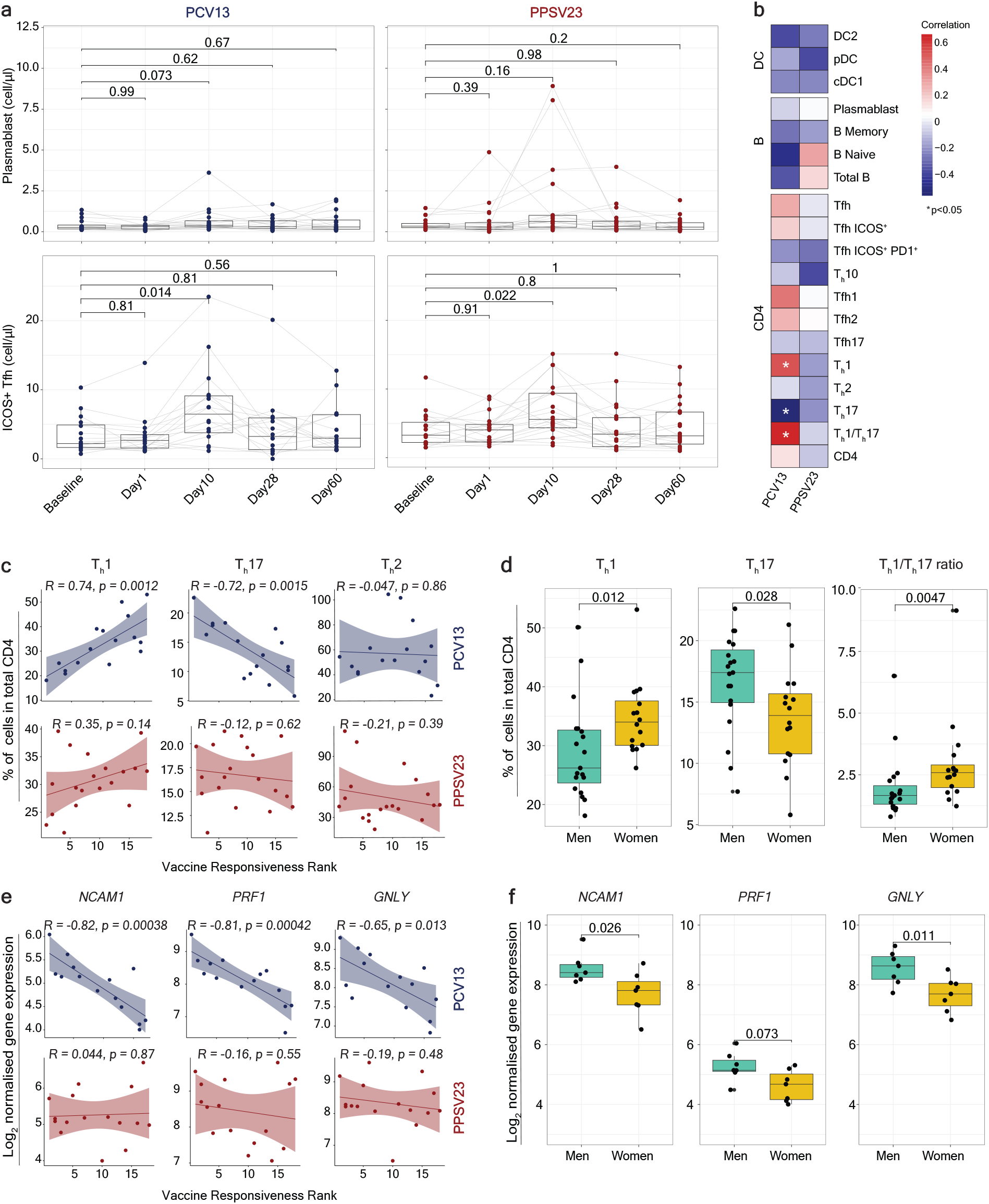
Baseline T_h_1/T_h_17 cell ratio and cytotoxic gene expression are predictive of PCV13 vaccine responsiveness Rank. **a**) Longitudinal analysis of the absolute number of plasmablasts(cells/ul) (top panel) and ICOS^+^ Tfh(cells/ul) cells (bottom panel) among the memory CD4+ T cell population in response to PCV13 and PPSV23. **b**) Correlation analysis between the absolute number of the different cell types (DCs, B and CD4 subsets) analyzed in whole blood and Ranks. **c**) Correlation analysis between Ranks and frequency of T_h_1, T_h_17, and T_h_1/T_h_17 ratio evaluated at baseline. **d**) Sex differences in the frequency of T_h_1, T_h_17 and T_h_1/T_h_17 at baseline. T_h_1 and T_h_17 cell frequency were calculated relative to the total CD4 Tcell count. **e**) Correlation analysis between baseline expression of cytotoxic genes (*NCAM1*, *GNLY*, and PRF1) and PCV13 vaccine responsiveness Rank (top panel). Correlation analysis between baseline expression of cytotoxic genes (*NCAM1*, *GNLY*, and PRF1) and PPSV23 vaccine responsiveness Rank (bottom panel). **f**) Sex differences in the expression of *NCAM1*, *PRF1* and *GNLY* at baseline. The Wilcoxon matched-pairs signed-rank test was used to compare the absolute numbers of plasmablasts and ICOS^+^ Tfh longitudinally (a). Correlation analysis was performed using the Pearson correlation metric (b,c,e).

To uncover baseline cellular predictors, we performed Pearson correlation analyses between the pre-vaccination absolute cell numbers for B, T, and DC subsets and vaccine responsiveness (rank) for each vaccine. None of the profiled cell types were significantly associated with PPSV23 rank (**Fig. 3b**). For PCV13, we detected a significant positive association with T_h_1 cell frequency (R=0.74, p=0.0012), and a negative association with pro-inflammatory T_h_17 cell frequency (R=-0.72, p=0.0015) (**Fig. 3b-c**, see Fig. s5a-b for cell numbers). The ratio of T_h_1 to T_h_17 cell frequency was the strongest predictor of responsiveness to PCV13 (R=0.63, p=0.0089) and was specific to this vaccine (**Fig. 3b, Fig. s5a**). There was no statistically significant difference in Tfh frequency between responders and non-responders, although Tfh1 showed a positive and Tfh17 showed a negative association with PCV13 responsiveness (**Fig. 3b**). We also observed sex differences in T_h_1 and T_h_17 cell frequency, women had significantly higher frequency of T_h_1 (p=0.012) and significantly lower frequency of T_h_17 (p=0.028) compared to men (**Fig. 3d, Fig. s5c**). Women with higher ratio of T_h_1/T_h_17 cells tended to mount stronger responses to PCV13 (**Fig. s5c**). These two cell types and their ratio was not significantly associated with age for either sex (**Fig. s5d**). Together, these data established that individuals with higher frequency of T_h_1 cells and lower frequency of T_h_17 cells (hence higher ratio of T_h_1 to T_h_17) prior to vaccination mount stronger responses to T-dependent PCV13 vaccine.

### The baseline expression of cytotoxicity associated genes (CYTOX) is negatively associated with T-dependent PCV13 vaccine responses

To identify baseline transcriptional signatures associated with responsiveness to both vaccines, we conducted Weighted Gene Co-expression Network Analysis (WGCNA)^(29)^. Analyses of pre-vaccination PBMC RNA-seq data for both cohorts (n=30) yielded 33 gene modules containing 70 to 1523 genes, which were further studied for any association with vaccine responsiveness (PCV13-Rank, PPSV23-Rank), sex, age and T_h_1/T_h_17 ratio in each vaccine cohort. Out of 33 modules, only one module (midnight blue containing 260 genes) was significantly (p=0.02) associated with reduced PCV13 responses, an association that was not detected for PPSV23 (p=0.53) (**Fig. s6a, Supplementary Table S5a**). Genes in this midnight blue module were significantly enriched in “Natural killer cell mediated cell cytotoxicity” KEGG pathway (p=0.000003639) and included *NCAM1,* the marker gene for NK cells and several transcripts (*GNLY, PRF1, and GZMB*) encoding for cytotoxic effector functions^(30–32)^ (**Supplementary Table S6a**). This midnight blue cytotoxicity-associated gene set is hereafter referred to as the CYTOX signature. Baseline expression levels of CYTOX genes in PBMCs was negatively associated with responsiveness to T-dependent PCV13 (e.g., for *NCAM1* R= -0.82 and p= 0.00038), but not to PPSV23 (**Fig. 3e**). Expression levels of the top CYTOX genes increased with age in this cohort albeit not significantly (**Fig. s6b**); age-related upregulation of these genes was also observed in a larger cohort based on PBMC RNA-seq data from 75 donors (n= 41 men, 34 women) (p <0.05) ^(33)^ (**Fig. s6c**). In addition, PBMCs from men on average had higher expression of these CYTOX genes than women in both cohorts (**Fig. 3f, s6d**). A different module (dark green containing 134 genes) was specifically associated with reduced PPSV23 responses (p=0.05 for PPSV23 and p=0.67 for PCV13) (**Fig. s6a,e**, **Supplementary Table S6b)**. This module included genes associated with cell cycle and transcriptional regulation (*ANGEL2, MRE11, MTERF1, NUP107, YTHDC2*), however, there was not a significant association with any immune cell type or function for this gene set. In sum, WGCNA of the baseline PBMC RNA-seq data revealed two novel and distinct gene sets that are significantly associated with reduced responsiveness to PCV13 and PPSV23 vaccines.

### Increased frequency and increased cytotoxic gene expression in CD16^+^ NK cells are associated with reduced PCV13 vaccine responsiveness

To identify cells bearing the baseline CYTOX signature, we generated single cell RNA-seq (scRNA-seq) data from pre-vaccination PBMCs of 11 PCV13 donors: 6 top responders referred to as responders (R) and 5 bottom responders referred to as non-responders (NR), resulting in 52,702 cells after quality control (**Fig. 4a, Supplementary Table S7**). Clustering of PBMCs generated 24 clusters, which were then annotated into subsets of monocytes (CD14, CD16), dendritic cells (monoDCs, cDC1, cDC2), B cells (naive, memory, switched memory and age-associated B (ABCs), and plasma cells), CD4^+^ T cells (naive, memory, Treg, cytotoxic CD4^+^ T cells), CD8^+^ T cells (naive, memory (GZMK^+^, TEMRA, MAIT, gamma delta)) and NK cells (CD56^bright^CD16^-^, CD56^dim^CD16^+^) using established markers^(31, 32)^ (**Fig. 4a, Fig. s7a,b**). PBMC cell composition analysis revealed that only two cell subsets were significantly different between PCV13 responders and non- responders (**Fig. 4b**, **s8a**); responders had more naive CD8^+^ T cells (p=0.03) and fewer CD16^+^ NK cells (p=0.03) (**Fig. 4b**, **s8a**).

**Figure 4:**
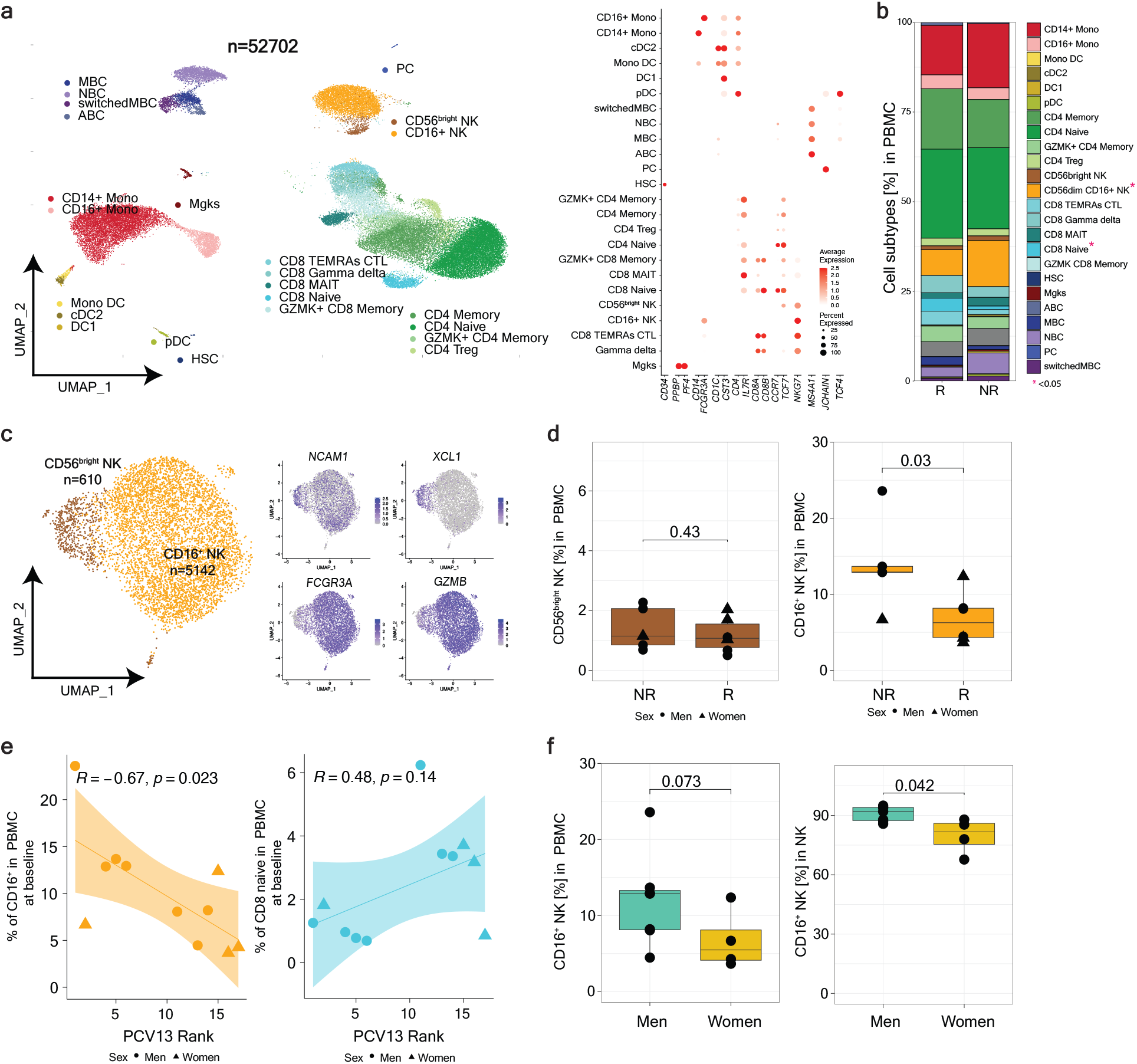
CD16^+^ NK cell frequency in PBMCs is negatively associated with PCV13 vaccine responses. **a)** Uniform manifold approximation and projection (UMAP) of PBMCs from 11 PCV13 donors (responders (R)=6 and non-responders (NR)= 5) showing 24 clusters from 52,702 cells, colored by immune cell type. Immune subsets were identified in a supervised manner. Lineage markers are shown in the dot plot. **b**) Stacked bar plot of immune cell frequency in R and NR. The cell types with significant differences in their frequency between R and NR are marked with a red star (p-value < 0.05). **c**) UMAP of NK subsets with feature plot showing the expression *NCAM1*, *XCL1*, *FCGR3A* and *GZMB* in blue, highlighting the two NK populations: CD56^dim^ CD16^+^ NK and CD56^bright^ NK. **d**) Boxplots of CD16^+^ NK and CD56^bright^ NK frequency in R and NR. **e**) Correlation analysis between PCV13 Rank and pre-vaccination frequency of CD16^+^ NK and CD8+ Naïve T cells. **f**) Sex differences in the pre-vaccination percentages CD16^+^ NK in total PBMC and in total NK. The Wilcoxon Rank sum test was used to compare cell percentages between R and NR (b, d), and CD16^+^ NK percentages between men and women (f). Correlation was computed using the Pearson correlation metric (e).

Blood NK cells include immature CD56^bright^ CD16^-^ NK cells that produce the chemokine XCL1 and the mature highly cytotoxic CD56^dim^CD16^+^ cells, both of which was captured in the scRNA-seq data (**Fig. 4c**)^(34)^. However, only the CD16^+^ NK cells were significantly associated with the vaccine response; where non-responders had higher frequencies of these cells (p=0.03 for CD16^+^ NK cells, p=0.43 for CD56^bright^ NK cells) (**Fig. 4d**). We also correlated the responsiveness for each donor (i.e., rank) with the frequency of NK cells and other immune cell subsets. Out of all PBMC subsets, only the frequency of CD16^+^ NK cells was significantly associated with PCV13 responsiveness (R=-0.67, p=0.023) (**Fig. 4e, s8b**). As expected from their stronger PCV13 responses (**Fig. 1f**), women had lower CD16^+^ NK cell frequency compared to men (p=0.042 for comparisons within the NK lineage) (**Fig. 4f**) ^(35)^. Together these data establish that the increased frequency of CD16^+^ NK cells bear the CYTOX signature that is associated with reduced PCV13 responses.

Differential gene expression analyses between CD16^+^ NK cells of responders and non-responders, revealed 38 differentially expressed genes (FDR 5%; **Fig. 5a**, **Supplementary Table S8**). CD16^+^ NK cells from non-responders had higher expression of cytotoxic genes (*GNLY, GZMH*) and *IFNG*, indicating a more ‘cytotoxic’ and ‘activated’ phenotype. In addition, non-responder CD16^+^ NK cells expressed higher levels of the inhibitory receptor *KLRC2 (NKG2C)*, suggesting a more advanced maturation stage^(34, 36)^. Non-responder CD16^+^ NK cells also overexpressed Amphiregulin (*AREG*), a ligand for the epidermal growth factor receptor (EGFR), that is upregulated upon inflammation and mitochondrial stress in immune cells^(37)^. Though CMV can modulate the NK cell phenotypes^(38)^, CMV positivity (10 out of 39 participants) was not associated with responsiveness to either vaccine (**Fig. 5b**). These data suggest that CD16^+^ NK cells from PCV13 non-responders had a more cytotoxic and activated transcriptional phenotype compared to the same cells from responders.

**Figure 5:**
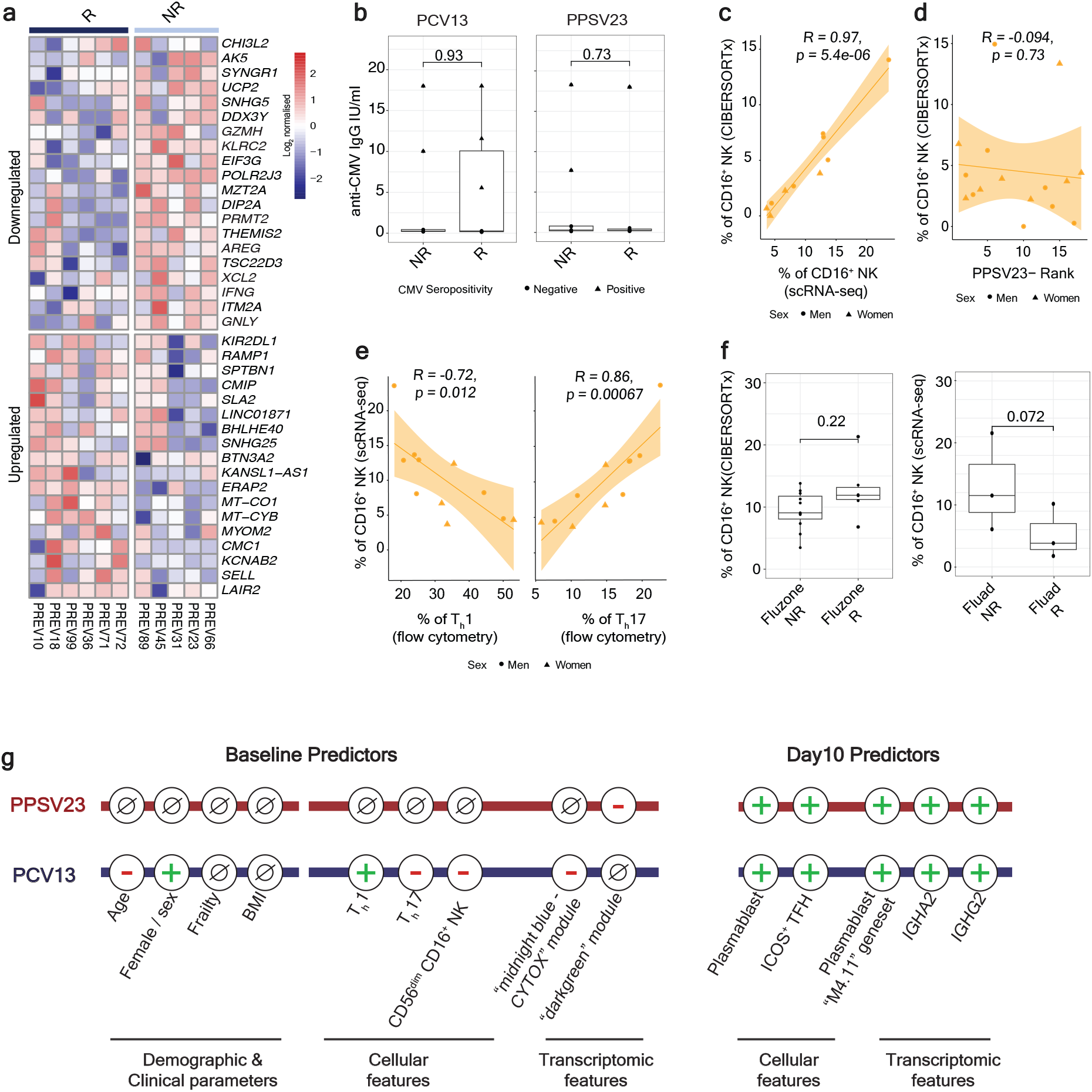
Increased cytotoxicity in the CD16^+^ NK cells of PCV13 non-responders. **a**) Heatmap of the differentially expressed genes in the CD16^+^ NK cells of PCV13 responders (R) and non-responders (NR) at baseline, using the normalized expression values from the scRNA-seq pseudobulk analysis. **b**) Boxplots comparing anti-CMV IgG titers between R and NR for PCV13 (left) and PPSV23 (right). **c)** Correlation analysis of pre-vaccination CD16^+^ NK percentages estimated by scRNA-seq and CIBERSORTx. **d)** Correlation analysis of CIBERSORTx based estimates of CD16^+^ NK and PPSV23-Rank (n=16) at baseline. **e**) Correlation analysis CD16^+^ NK percentages determined by scRNA-seq, and T_h_1 and T_h_17 percentages determined using flow cytometric analysis, at baseline. **f)** Boxplots of pre-vaccination CD16^+^ NK percentages in FluAd-R(n=3) and NR(n=3), and Fluzone TIV R(n=5) and NR(n=11). **g)** Summary schema showing the demographic, clinical, cellular and transcriptomic parameters associated with PCV13 and PPSV23 vaccine responsiveness at baseline and Day 10. The Wilcoxon Rank sum test was used to compare the mean of anti-CMV IgG titers between R and NR of PCV13 and PPSV13 donors (b), pre-vaccination CD16^+^ NK percentages in FluAd-R and NR, and Fluzone R and NR (f). Correlation analysis was computed using Pearson correlation metric (c, d, e).

### CD16^+^ NK cell frequency is not associated with PPSV23 vaccine responsiveness

To establish whether CD16^+^ NK cells are also associated with responsiveness to PPSV23, we used CIBERTSORTx^(39)^, a machine learning framework that can infer cell type frequency from bulk data by utilizing cell-type-specific expression patterns inferred from scRNA-seq data. We trained the CIBERTSORTx model on cell-specific signatures from PBMC scRNA-seq data (**Fig. 4a**) and confirmed its efficacy to infer CD16^+^ NK cell frequencies for 11 donors for whom we have both single cell and bulk PBMC RNA-seq data (R=0.97, p=5.4e-06) (**Fig. 5c**, **Fig s9a** for other cell types). Next, we inferred the frequency of CD16^+^ NK cells from the bulk RNA-seq data for all profiled donors (14 PCV13, 16 PPSV23). In the PCV13 cohort, inferred CD16^+^ NK frequency significantly correlated with reduced responsiveness as expected (R=-0.69, p=0.0067) (**Fig. s9b**). In contrast in the PPSV23 cohort, the CD16^+^ NK frequency was not associated with vaccine responsiveness (R=-0.094, p=0.73) (**Fig. 5d, s9b**). Together with the lack of association of CYTOX genes with PPSV23 responsiveness (**Fig. 3e**), these analyses established that CD16^+^ NK cells play a role only for the conjugated PCV13 vaccine responses. Notably, donors with higher CD16^+^ NK frequency also had higher T_h_17 frequency (R=0.86, p=0.00067) and lower T_h_1 frequency (R=-0.72, p=0.012) (**Fig. 5e**). The association between T_h_ subsets and NK cell frequencies was specific to CD16^+^ NK cells (**Fig. s9c**).

To study whether CD16^+^ NK cells are critical for responsiveness to other vaccines in older adults, we reanalyzed bulk PBMC RNA-seq data from a Fluzone study (n=16, 65+ years old)^(40)^. CIBERSORTx-inferred CD16^+^ NK cell frequencies did not show significant differences between responders and non-responders for Fluzone (p=0.22, **Fig. 5f**). However, re-analyses of scRNA-seq data from a recent FluAd study (n=6, 65 years old)^(41)^, showed that CD16^+^ NK cell frequency was higher in non-responders compared to responders despite the small cohort size (3 responders, 3 non-responders) (p=0.072, **Fig. 5f**). FluAd is the first FDA approved adjuvanted (MF59) influenza vaccine approved in the US for the use among older adults to increase their responsiveness to the influenza vaccine^(42–44)^. Together, these data suggest that the novel CD16^+^ NK phenotype and the CYTOX signature is specifically associated to conjugated pneumococcal vaccine PCV13, providing an opportunity to stratify the population for alternative pneumococcal vaccines. Analyses of data from different influenza vaccine studies showed that this signature might be predictive of responses for other vaccines as well (e.g., FluAd) and might provide an effective strategy for precision vaccinology among older adults. Associations of this signature with other vaccines should be studied further in larger cohorts.

## DISCUSSION

Older adults are at high risk for morbidity and mortality due to infectious diseases^(45)^ including those caused by *Streptococcus pneumoniae* (Pneumococcus). Despite the availability of two types of FDA approved vaccines for pneumococcus, responsiveness of older adults to these vaccines remains poorly understood. To fill this knowledge gap, we recruited 39 older adults (60+ years old) and vaccinated them in 2017-2018 seasons with the two available vaccines at the time: PPSV23 (Pneumovax®), and conjugated PCV13 (Prevnar®)^(46, 47)^. We used diverse assays including RNA-seq (bulk and single cell), flow cytometry, and opsonophagocytosis assays (OPA). In summary, we uncovered 1) strong antibody responses to both vaccines and transcriptional activation of plasmablast genes at day 10; 2) increased ICOS^+^ Tfh cells at day 10 for both vaccines; 3) an activated immune phenotype composed of increased T_h_17, reduced T_h_1, and increased CD16^+^ NK cell frequencies that is specifically associated with reduced PCV13 responses and more frequently observed in men compared to women; and 4) a distinct baseline gene expression module that is associated with PPSV23 responses (**Fig. 5g**).

Both vaccines induced strong antibody responses, though PCV13 induced higher increases in OPA titer levels than PPSV23, in line with a previous study that enrolled younger (50- to 64-year-old) donors^(48)^. At the serotype level, both vaccines induced strong responses to the 12 shared serotypes. Although, PPSV23 lacks serotype 6A, some donors vaccinated with PPSV23 had increased antibody responses to this serotype, potentially due to the cross-reactivity with serotype 6B^(20)^. In the longitudinal RNA-seq data, the strongest signal was the upregulation of plasmablast genes – particularly immunoglobulins – at day 10. Free polysaccharides, such as those present in PPSV23, induce T cell-independent responses and are associated with the induction of the IgG2 isotype^(46, 47)^. In alignment, both vaccines predominantly induced the expression of the heavy chain constant region genes encoding IgG2, and IgA2 isotypes^(49)^, distinct from the influenza vaccine responses (IgG1, IgG3). A recent systems serology study on PCV13 and PSSV23 (n=20 per vaccine arm, 60-64 years old)^(25)^, showed that there is increased abundance of these isotypes 30 days after the vaccination for both vaccines. Despite the strong transcriptional upregulation of plasmablast genes at day 10, increases in the numbers of circulating plasmablasts at this timepoint were marginal. It is possible that 10 days post-vaccination was not the ideal timepoint to detect the peak plasmablast responses or the responses were weaker in older adult populations. Indeed, in a previous study we showed that antigen specific (AIM^+^) Tfh cells are only expanded in younger donors (n=5) and not observed in older donors (n=5) upon PCV13 vaccination^(6)^.

The release of plasma cells into circulation is intimately linked to the frequencies of activated Tfh cells that positively correlate with serological responses to influenza vaccine^(28, 50)^ and are affected by aging^(51)^. For pneumococcal vaccines, although there was a significant expansion of ICOS^+^ Tfh cells at day 10, this expansion did not significantly correlate with antibody responses, which could be due to age of our donors or the timing of our Tfh measurement (day 10 versus day 7). In our study, the expansion of ICOS^+^ Tfh cells in response to PPSV23 – at comparable levels to PCV13 – suggests that PPSV23 might also induce T cell immunity, forcing us to reconsider the human immune responses to T cell-independent vaccines. Surprisingly, having a lower frequency of T_h_1 cells and higher frequency of T_h_17 cells at baseline was associated with weaker responses to PCV13. Age-dependent increases in T_h_17 cells or increased production of IL-17 have been observed in previous studies^(52, 53)^. In this cohort, the frequency of these cell types were not significantly associated with age, however, there was a significant sex association. Women had higher T_h_1 and lower T_h_17 cell frequency compared to age-matched men, which explained their stronger responses to PCV13. It is noteworthy that the proinflammatory molecules IL6 and IL1β that promote T_h_17 cell differentiation^(54)^ are also linked to systemic chronic inflammation with aging (i.e., inflammaging)^(55–60)^. Previously, we uncovered an accelerated aging phenotype in men compared to women^(33)^, which might contribute to the observed sex differences in PCV13 responses. This association needs to be further investigated in larger cohorts. In contrast to our earlier data generated from healthy young adults^(16)^, and despite the association of baseline CD16^+^ NK cells with PCV13 vaccine responses, we did not detect an innate response at day 1 for either PPSV23 or PCV13. This might be due to the already elevated levels of baseline inflammation^(57-59^, ^61-64)^ in older individuals, or to an age-specific altered response to these vaccines.

Baseline bulk RNA-seq data revealed a cytotoxicity-associated gene set (CYTOX) that predicted reduced responses to conjugated PCV13. ScRNA-seq data suggested that this CYTOX signature specifically stemmed from mature CD16^+^ NK cells that are more cytotoxic than immature CD56^bright^ NK cells and that have the antibody dependent cellular cytotoxicity (ADCC) functions^(30)^. Although ADCC-proficient antibody production is a key correlate of vaccine responses, including for bacterial pneumonia vaccines^(25)^, several studies have reported that activated NK cells might be detrimental for vaccine responses. For the yellow fever vaccine YF-17D (n=50 young adults), the frequency of activated NK cells at day 7, was linked to reduced neutralizing antibody levels, though there was no report of an association between baseline NK cell levels and YF-17D responses^(65)^. For malaria vaccines (n=46), blood transcriptional modules associated with NK cells displayed strong negative correlation with antibody response and protection at multiple time points (Days 2, 28, 29, 56)^(66)^.

scRNA-seq data revealed that CD16^+^ NK cells from non-responders expressed cytotoxicity related genes at higher levels than those from responders. These cytotoxic molecules and some NK receptors are also upregulated with age in CD8^+^ TEMRA cells^(67)^. Although we captured the TEMRA population in the scRNA-seq data, these cells were not associated with PCV13 responses. Interestingly, CD16^+^ NK cell frequency significantly correlated with Th subsets that are associated to PCV13 responses. Donors who had higher frequency of CD16^+^ NK cells had higher frequency of T_h_17 cells and lower frequency of T_h_1 cells. Whether this reflects a parallel co-regulation of these three subsets in older donors or whether the CD16^+^ NK cells contribute to the imbalance of T_h_1 and T_h_17 remains to be established. Studies in mice have indicated that NK cells can kill Th and Tfh cells in certain viral infection settings, resulting in worse CTL responses, reduced GC formation, B cell affinity maturation, and neutralizing antibody titers^(68–72)^. Indeed, NK cell depletion before vaccination increased vaccine induced antibody responses in mice^(71)^. Whether this applies to humans remains to be determined.

This is the first precision vaccinology study to examine how older adults respond to two available types of pneumococcal vaccines. Remarkably, the baseline predictors for the two vaccines were distinct and independent from each other, despite the fact that both vaccines include the same polysaccharidic antigens. Our study provides a framework for the potential future point-of-care clinical stratification of older adults for the administration of individually selected pneumococcal vaccines, with potential implications for other vaccines as well. Simple assays to measure *NCAM1* expression or CD16^+^ NK frequency in the blood could be established to assess baseline cytotoxicity levels. Based on our findings, older adults with lower cytotoxicity are likely to mount strong responses to the PCV13 vaccine. Since this association was vaccine-specific, older adults with high cytotoxicity are more likely to benefit from the PPSV23 vaccine. Our data also showed the importance of taking into consideration biological sex while administering these vaccines; women had stronger responses to T-dependent PCV13 since they were less likely to exhibit the CYTOX signature compared to age-matched men. These findings might also give rise to novel intervention strategies to change the baseline immune phenotypes to alter the outcome of vaccine responses^(13)^, e.g., by temporarily blocking the immunosuppressive or cytolytic functions of CD16^+^ NK cells prior to vaccination^(73)^.

## Data availability

All PBMC RNA-seq samples used in this study can be found at dbGAP accession code: phs002361.v2.p1. https://www.ncbi.nlm.nih.gov/projects/gapprev/gap/cgi-bin/study.cgi?study_id=phs002361.v2.p1

## Author contributions

J.B. designed the study and raised the funds. J.B., G.A.K., D.U. co-supervised the study. G.A.K. coordinated human subject studies and sample collection. L.K.P. developed recruitment strategies and facilitated blood collection. M.H.N. coordinated and generated OPA titer data. R.M., R.R. generated flow cytometry, RNA-seq, and CMV data. S.R., F.E.D, and O.E.K analyzed the data. D.U., F.E.D, J.B., S.R. wrote the paper. V.P., K.P, S.P., D.C. helped with data interpretation and edited the manuscript. All authors revised the manuscript and figures prior to submission.

## Data Availability

All data produced in the present study are available upon reasonable request to the authors.

## Acknowledgements

We thank Taneli Helenius for aid in scientific writing, Magalie Collet for help with dbGAP data upload, research staff in the UConn Center on Aging for their help in recruitment and sample collection, Diane Luo and Paul Gabriel from JAX Genomic Technologies and Single Cell cores for their help with generating the sequencing data. JAX single cell service is supported in part by the JAX Cancer Center (P30 CA034196). We thank members of the Ucar lab for critical feedback during the progress of the study. This study was made possible by generous financial support of the National Institutes of Health (NIH) grants under award number - R01AG052608 (to J.B.), U01AI165452, P30AG067988 Older Americans Independence Pepper Center (to G.A.K., R.M., D.U.) and R01AI142086 (to DU, GAK), and JAX Cancer Center Brooks Scholarship (to S.R.). Opinions, interpretations, conclusions, and recommendations are solely the responsibility of the authors and do not necessarily represent the official views of the National Institutes of Health (NIH).

## Supplementary table legends

**Table S1: Donor Demographics.** Donor level details such as vaccine received, age (in years), sex, race, ethnicity, BMI, CMV seropositivity, and Frailty index, and Cohort level demographics summary are provided. P-values were determined using the two-sided t-test for continuous variables, considering each donor parameter individually.

**Table S2: Functional Antibody Response to PCV13, and PPSV23.** OPA titers at Day -7 (baseline), Day 28, and the fold difference in their antibody titers (D28/baseline) for PCV13 and PPSV23, respectively. Vaccine responsiveness scores such as Strength, Extent, Rank, and maxRBA, which assess the response to PCV13, and PPSV23 are provided.

**Table S3: Gene expression changes in PBMC upon PCV13, and PPSV23 vaccination.** Gene expression changes at Day 1, Day 10 and Day 60 upon PCV13 in comparison to baseline, as well as the gene expression changes at Day 1, Day 10 and Day 60 upon PPSV23 in comparison to baseline. Genes with log_2_ FC > ± 0.585 (i.e., 1.5) and FDR pvalue < 0.05 were considered to be significantly differentially expressed.

**Table S4: Longitudinal Profiling of DCs, B Cells, and CD4+ T Cell Subsets in PCV13 and PPSV23 donors.** Absolute numbers of immune cell subsets at baseline, Day 1, Day 10, and Day 60. The markers used for profiling these immune subsets are provided in the table.

**Table S5: Gene Co-expression-based modules inferred using WGCNA.** The table presents the thirty-three modules identified by WGCNA, along with their correlation with metadata such as PCV13 Rank, PPSV23 Rank, sex, age, and T_h_1/T_h_17 ratio. Correlation analysis was performed for PCV13 and PPSV23 donors separately using Pearson correlation as a correlation metric.

**Table S6: PCV13–associated Midnightblue (i.e., CYTOX) and PPSV23–associated Darkgreen module**. Genes and pathways enriched in midnightblue and darkgreen module. The Midnightblue module is significantly and negatively correlated to PCV13, while Darkgreen module is significantly and negatively correlated to PPSV23.

**Table S7: Number of cells before and after preprocessing filters across 11 PCV13 donors (6 Responders and 5 Non-responders) at baseline.** Preprocessing filters include doublet removal, and other steps described in methods.

**Table S8:** Differentially expressed genes in CD56^dim^ CD16^+^ NK cells.

Genes with log_2_ FC > ± 0.25, FDR pvalue < 0.05 and expressed in at least 10% of the cells in both groups (R, NR) were considered to be significantly differentially expressed.

## METHODS

### Human subject recruitment

All studies were conducted following approval by the Institutional Review Board of UConn Health Center (IRB Number: 16-071J-1) and registration on ClinicalTrials.gov (NCT03104075), and written informed consent was obtained from each volunteer. The volunteer identifiers were decoded and used for the study. Blood samples were obtained from 39 healthy volunteers residing in the Greater Hartford, CT, USA region recruited by the UConn Center on Aging Recruitment and Community Outreach Research Core (http://health.uconn.edu/aging/research/research-cores/). Volunteers were vaccinated from May to early fall to avoid overlap with potentially confounding seasonal peak periods of influenza vaccination and infection. None of the participants reported a history of a previous pneumonia. Recruitment criteria were selected to identify individuals who are experiencing “usual healthy” aging and are thus representative of the average or typical normal health status of the local population within the corresponding age groups. Selecting this type of cohort is in keeping with the 2019 NIH Policy on Inclusion Across the Lifespan (NOT-98-024), increasing the generalizability of our studies and the likelihood that these findings can be translated to the general population. Inclusion criteria included age 60 years or older plus willingness to receive pneumococcal vaccination with Prevnar 13 (Wyeth/ Pfizer) or Pneumovax 23 (Merck) via random assignment. Exclusion criteria included having previously received Prevnar 13 or Pneumovax 23 vaccines or a history of adverse reactions to these or any diphtheria toxoid-containing vaccine. Individuals were also excluded if they had a recent fever, received a Zostavax (shingles vaccine) in the previous 4 weeks on in the presence of significant potentially confounding co-morbidities such as diabetes mellitus, active malignancy or recurrence in the last 5 years, congestive heart failure, cardiovascular disease unstable in the last 6 months, renal failure, impaired hepatic function, autoimmune diseases (e.g. rheumatoid arthritis, lupus, inflammatory bowel disease), recent trauma or surgery, HIV or other immunodeficiency, current substance, or alcohol abuse, as well use of medications known to alter the immune system (e.g., prednisone ≥ 10 mg or biological medications).

### Sample collection and PBMC isolation

Thirty-nine healthy participants aged over 60 received either Prevnar-13 (PCV13) or Pneumovax (PPSV23) vaccine at the University of Connecticut Health (UCH) at Farmington, CT. Under an IRB approved study, peripheral blood was collected at baseline (prior to vaccination, day -7) and after vaccination (days 1, 10, 28, and 60). PBMCs were isolated from peripheral blood samples collected in ACD tubes by using Lymphoprep gradient (StemCell Technologies) whereas serum was isolated from blood collected in red top collection tubes (BD Vacutainer). Both PBMCs and serum were cryopreserved prior to subsequent analytical steps.

### OPA titer measurements

MOPA was performed for 13 serotypes as described in^(17)^. We analyzed serotypes 1, 3, 4, 5, 6A, 6B, 7F, 9V, 14, 18C, 19A, 19F, and 23F. For this, frozen aliquots of target pneumococci were thawed, washed twice with Opsonization Buffer B (Hanks’ balanced salt solution (HBSS) with Mg/Ca, 0.1% gelatin, and 10% fetal bovine serum) by centrifugation (12,000 x g, 2 minutes), and diluted to the proper bacterial density (∼2 x 10^5^ cfu/ml of each serotype). Equal volumes of 4 bacterial suspensions that were chosen to be analyzed together, were pooled. All serum samples were incubated at 56°C for 30 minutes before serial dilutions in Opsonization Buffer B. Serially diluted serum (20 μl/well) was mixed with 10 μl of bacterial suspension in each well of round bottom 96-well plates.

After 30-minute incubation at RT with shaking at 700 rpm on mini orbital shaker (Bellco Biotechnolgy, Vineland, NJ) 10 μl of 3-4 weeks-old rabbit complement (PelFreeze Biologicals, Rogers, Arkansas) and 40 μl of HL60 cells (4 x 10^5^ cells) were added to each well. HL60 cells were differentiated to granulocytes by culturing in RPMI-1640 with 10% fetal bovine serum and 1 % L-glutamine and 0.8% dimethylformamide at a starting density of 4 x 10^5^ cells/ml for 5-6 days. Plates were incubated in a tissue culture incubator (37°C, 5% CO_2_) with shaking at 700 rpm. After a 45 minute incubation, plates were placed on ice for 10-15 minutes and an aliquot of the final reaction mixture (10 μl) was spotted onto 4 different THY agar plates (Todd-Hewitt broth with 0.5 % yeast extract and 1.5% agar). When the fluid was absorbed into the agar, an equal volume of an overlay agar (THY with 0.75% agar and 25 mg/L of TTC) containing one of the four antibiotics was applied to each THY agar plate. After an overnight incubation at 37°C, the number of bacterial colonies in the agar plates was enumerated. Opsonization titers were defined as the serum dilution that kills 50% of bacteria.

### OPA Titer Ranking Methods

Three different strategies were developed to quantify the antibody responses, namely: RANK (responsiveness in the cohort), STRENGTH (sum(log_2_)), and EXTENT (number of serotypes an individual develops strong responses). We used the dense ranking strategy to rank individuals in a cohort in terms of their responsiveness to the vaccine using their responses to all serotypes. First, each subject is dense ranked for each serotype according to their OPA titer levels: baseline ranking or fold change (post/pre) ranking. Subjects with equal titer (or FC) levels receive the same ranking number, and the next subject receives the immediately following ranking number. This generates a rank for each subject for each serotype, which are then summed up to find an overall score for the individual. Next, we applied dense-ranking on these overall scores to appoint the final rank of each individual. In this measure, individuals with higher titer levels across many serotypes will rank higher than others, whereas individuals with lower titer levels across many serotypes will rank lower than others. This ranking strategy is robust to outliers (i.e., a subject with very high titer levels only for one serotype) and is based on a multivariate approach (using all 13 serotypes) to quantify vaccine responsiveness of individuals. We applied the ranking strategy to both baseline titer levels (Rank(baseline)) and to changes between pre- and post-vaccination (Rank (foldchange)). sum(log_2_) (i.e. STRENGTH) of the OPA titer levels is the sum of changes in OPA titer levels for all serotypes. It is used to explain the dynamics of the baseline and post-vaccination changes per sample. Individuals with a high score have stronger responses than individuals with lower scores. In the literature, 8-fold change of a given OPA titer level is accepted as the response to a specific serotype, and the extent of the response strategy was built on this fact^(20)^. EXTENT is defined as the number of serotypes (out of 13) which responds to vaccination, and it is calculated per individual. Individuals who develop significant responses across many serotypes will have higher values for the extent score. These measures have unique advantages, but they also agree in many respects especially in determining responsiveness of individuals (**Fig. s1a,b**). maxRBA method^(22)^ corrects for the dependence of baseline antibody titers by modeling fold change as an exponential function of pre-vaccination levels and factoring in the maximum residual across all serotypes. To calculate the baseline-adjusted fold change for 13 serotypes, the maxRBA function (titer package in R) with scoreFun set to “sum” was employed. The adjusted fold change was then utilized to rank individuals using a dense ranking approach.

### Determination of CMV seropositivity

CMV specific IgG antibodies to human CMV were measured in serum samples of participants by enzyme-linked immunosorbent assay (ELISA) using the CMV immunoglobulin (IgG) Enzyme Immunoassay kit (Aviva Systems Biology). Serum samples were thawed, centrifuged to 6,000xg for 1 minute and samples and controls dilution was prepared according to the manufacturer’s recommendations. Calculation of results was done with a nonlinear regression curve fit based on the controls provided by the vendor. Seropositivity was defined as a concentration of CMV-specific IgG of ≥1 IU/mL.

### Flow cytometry data generation and analyses

Fluorescent-labeled antibody cocktails for different cell-surface staining panels were pre- mixed in BD Horizon Brilliant Stain Buffer (BD Biosciences) 10 minutes before staining. Antibody cocktails were added over 100 μL aliquots of anticoagulated whole blood in a 5 ml FACS tube within 60 minutes of blood collection. Samples were incubated for 15 minutes at room temperature then lysed and fixed with 2 ml of 1x FACS lysing solution (BD Biosciences) for 8 minutes at room temperature. The lysed samples were washed twice to remove the unbound antibodies, lysed RBCs, and platelets and finally resuspended in 250 μL of PBS to which 50 μL of count beads suspension (Count Bright Absolute Counting Beads, Thermo Fisher) were added for the detection of absolute cell counts. For the analysis of CD4+ T-cell compartment, cells were stained with fluorochrome-labeled antibodies to the following surface markers: CD3 (clone UCHT1, BioLegend), CD4 (clone OKT4, BioLegend), CD183 (clone GO25H7, BioLegend), CD196 (clone 11A9, BD Biosciences), CD185 (clone J252D4, BioLegend), CD279 (clone MIH4, BD Biosciences), and CD45RA (clone 3H4, Beckman-Coulter). For the analysis of DC compartment, cells were stained with fluorochrome-labeled antibodies to the following surface markers: Lineage cocktail (Lin1) (CD3, clone SK7; CD16, clone 3G8; CD19, clone SJ25C1; CD14, clone MΦP9; CD20, clone L27; CD56, clone NCAM16.2; BD Biosciences), HLA-DR (clone LN3, ThermoFisher), CD11c (clone B-ly6, BD Biosciences), CD1c (clone L161, BioLegend), CD141 (clone AD5-14H12, Miltenyi Biotec), CD303 (clone AC144, Miltenyi Biotec), CD86 (clone IT2.2, BioLegend), and CD40 (clone 5C3, BD Biosciences). For the analysis of B-cell compartment, cells were stained with fluorochrome-labeled antibodies to the following surface markers: CD3 (clone UCHT1, BD Biosciences), CD19 (clone J3-119, Beckman Coulter), IgD (clone IA6-2, BD Biosciences), CD27 (clone M-T271, BD Biosciences), CD20 (clone 2H7, BioLegend), CD86 (clone IT2.2, BioLegend), CD38 (clone HIT2, BioLegend), and CD138 (clone MI15, BD Biosciences). The stained cells were acquired with LSR Fortessa X-20 (BD) and analyzed with FlowJo software (BD). For each vaccine, we assessed the statistical difference in the absolute number of each profiled cell type at Day 1, Day 10, Day 28, and Day 60, compared to baseline, using the Wilcoxon matched-pairs signed-rank test for high QC samples (n=35). We further associated the cellular composition at baseline with the (fold-change) RANKs of the donors (defined as ’RANK’ in Fig. 1b and 1c; see OPA Titer Ranking Methods) per cell type, including the Th1/Th17 ratio, using Pearson correlation. The correlation significance was calculated using the cor.test() function in R (v4.0.4).

### RNA-seq library generation

Total RNA was isolated from PBMCs using the Qiagen RNeasy Mini Kit (Qiagen) or Arcturus PicoPure (Life Technologies) kits following manufacturer’s protocols. During RNA isolation, DNase treatment was additionally performed using the RNase-free DNase set (Qiagen). RNA quality was checked using an Agilent 2100 Expert bioanalyzer (Agilent Technologies). RNA quality was reported as a score from 1 to 10, samples falling below threshold of 8.0 were omitted from the study. RNA-seq libraries were prepared with KAPA mRNA HyperPrep kit (Roche) according to manufacturer’s instruction. Poly (A) RNA was isolated from 200ng total RNA using oligo-dT magnetic beads and then fragmented at 85°C for 6 minutes, targeting 250-300 bp-range fragments. Fragmented RNA was reverse transcribed by incubating it for 10 minutes at 25°C followed by 15 minutes at 42°C and finally inactivated for 15 minutes at 70°C. This was followed by second strand synthesis and A-tailing at 16°C for 30mins and then at 62°C for 10 min. A-tailed, double stranded cDNA fragments were ligated with Illumina unique adapters (Illumina). Adapter-ligated DNA was purified using AMPure XP beads and amplified by 10 cycles of PCR amplification. The final library was cleaned up using AMPure XP beads. Quantification of libraries was performed using real-time qPCR (Thermo Fisher). Sequencing was performed on Illumina NovaSeq platform generating paired end reads of 150bp.Cyropreserved PBMCs are used for gene expression profiling. Libraries are generated using both KAPA mRNA Stranded and KAPA Hyper Prep protocols. RNA-seq libraries are sequenced using NovaSeq (2x150 bp paired-end reads) to obtain ∼100 million reads/sample.

### RNA-seq pre-processing

These RNA-seq samples were processed using the in-house pipeline at The Jackson Laboratory. Raw BCL files generated by the sequencer were converted to FASTQ files and sent to an in-house bioinformatics team for processing. CASAVA was used to generate FASTQ files for RNAseq data. Trimmomatic (version 0.33) was used to remove adapters, leading and trailing low-quality bases. Reads less than 36 bases long were dropped. Reads with more than 50% low-quality bases overall were filtered out. The remaining high-quality reads were then used for expression estimation. Alignment estimation of gene expression levels using the EM algorithm for paired-end read data was performed using RSEM (package version 1.2.12); default settings were used for alignment. RSEM uses bowtie2 as aligner to align the mapped reads against the hg38 reference genome. Data quality control was performed using Picard (version 1.95) and bamtools to obtain general alignment statistics from the bam file. Due to low quality of the baseline RNA-seq data PREV12 is excluded from downstream analyses.

### Gene expression data analyses

Gene expression values from PBMC RNA-seq data were converted into count-per-million (cpm) using edgeR R package (version 3.28.1)^(43)^. Genes with cpm values less than 0.5 across all samples were filtered out from the rest of the analyses. Differential gene expression analyses were conducted using generalized linear models (as implemented in the edgeR package) using the default TMM normalization. Significant genes were then determined according to p-values after Benjamini-Hochberg correction for multiple hypothesis testing. FDR p-value < 0.05 and log_2_ FC > ±0.585 were used to detect significant differences between time points and between vaccines. Plasma cell (M4.11) geneset was obtained from Obermoser et al. study^(16)^. Plasma cell activity score for a sample is given by the mean expression of the genes that comprises M4.11 geneset.

### Gene co-expression network analysis

To identify gene modules that are significantly associated with vaccine responsiveness measures, we utilized Weighted Gene Co-expression Analysis (WGCNA)^(29)^. Initially, we constructed a signed co-expression network using baseline transcriptomes (n=30) of PCV13 and PPSV23, and then identified gene modules using blockwiseModules() function in WGCNA package implemented in R (v4.0.4). Pearson correlation was used as the correlation metric with soft thresholding set to 18, deepSplit as 4, and mergeCutHeight set to 0.25. The number of genes in each module was restricted to range from 50 to 2000. Each module was represented by its first principal component of the gene expression values, also known as the module eigen gene. Next, we performed Pearson correlation analysis to examine the correlation between each of these gene module’s eigen genes and meta data such as PCV13-Rank, PPSV23-Rank, Age, Sex, and Th1/Th17 ratio. Modules with the Pearson correlation 0.5 and p-value < 0.05 were considered to be significant. Geneset enrichment analysis was conducted for the genes present in significantly correlated modules (Midnightblue module **Supplementary Table S6a;** Dark green module **Supplementary Table S6b**), using the hypergeometric test from the enrichR package in R^(74)^. The analysis included KEGG, Reactome, wikipathways, and BioPlanet as sources. Pathways with an FDR-corrected p-value of less than 0.05 were considered significant, and the Benjamini-Hochberg method was used to correct for multiple hypothesis testing.

### Sample processing and blood preparation for scRNA-seq

PBMCs were thawed quickly at 37°C and in DMEM supplemented with 10% FBS. Post-thawing, cells were washed and suspended in PBS containing 0.04% BSA and immediately processed as follows. Cell viability was assessed on a Countess II automated cell counter (ThermoFisher), and up to 12,000 cells (∼4,000 cells from each hashtagged sample) were loaded onto one lane of a 10 Genomics Chromium X. Single cell capture, barcoding and library preparation were performed using the 10x Chromium platform version 3.1 chemistry and according to the manufacturer’s protocol (#CG000388). cDNA and libraries were checked for quality on Agilent 4200 Tapestation and ThermoFisher Qubit Fluorometer, quantified by KAPA qPCR, and sequenced using 13% of an Illumina NovaSeq 6000 S4 v1.5 200 cycle flow cell lane, with a 28-10-10-90 asymmetric read configuration, targeting 6,000 barcoded cells with a minimum sequencing depth of 50,000 read pairs per cell.

### Single cell raw data processing

Illumina base call files for all libraries were converted to FASTQs using bcl2fastq v2.20.0.422 (Illumina) and FASTQ files associated with the gene expression libraries were aligned to the GRCh38 reference assembly with v32 annotations from GENCODE (10x Genomics GRCh38 reference 2020-A) using the version 6.1.2 Cell Ranger multi pipeline (10x Genomics). The generated gene - cell expression matrix was considered for the downstream analysis.

### Single cell gene-expression analysis

The 10X count matrices of PBMCs from 11 samples (6 PCV13-R and 5 PCV13-NR) were retrieved. Scrublet^(75)^ package in Python (V3.8) was used to estimate the doublet score for each sample with an estimated doublet set to 0.6, and cells identified as doublets were removed from downstream analyses. Then, a) cells with genes that were not detected in at least 3 cells, b) cells with mitochondrial reads greater than 20%, and c) cells with fewer than 500 or greater than 4000 features were excluded (**Supplementary Table S7**). Filtered gene expression matrices were merged and processed in accordance with the standard Seurat R workflow (version 4.0.3). We used the “NormalizeData’’ function to normalize the total number of reads in each individual cell to count-per-million (CPM). Next, the “FindVariableFeatures” function was used to select the 2,000 genes with the highest variance using the “vst” method. Then, the data were regressed against the percentage of mitochondrial genes and scaled to unit variance using the “ScaleData” function. Principal Component Analysis (PCA) was performed using the “RunPCA” function, followed by the “RunHarmony” function, from Harmony R package (version 1.0), to correct batch effects across samples. The first 50 Principal Components (PCs) were used in the “FindNeighbors” algorithm to construct the nearest-neighbor graph. Next, the Louvain modularity optimization algorithm in the “FindClusters” function was used to generate the clusters, while the resolution was set to 1.2. Then, the “RunUMAP” function was used to perform uniform manifold approximation and projection (UMAP). Finally, multiple rounds of marker identification, cell type annotation, and manual inspection and doublet removal, were performed to create the final UMAP. Genes that were differentially expressed between clusters were identified using the FindMarkers function in Seurat with the Wilcoxon rank-sum test.

Differences in cell frequency between PCV13 R and NR were calculated by computing the frequency of each immune cell type within PBMCs for each donor. The Wilcoxon rank-sum test was used to evaluate the difference in cell proportions between R and NR. Differentially expressed genes between PCV13 responders and non-responders for each cell type were identified from pseudo-bulked data using the edgeR package in R^(76)^. Genes with a log_2_ fold change greater than or less than ± 0.25, an FDR p-value less than 0.05, and expressed in at least 10% of the cells in both responder and non-responder groups were considered significantly differentially expressed.

CIBERSORTx analysis: We employed the CIBERSORTx algorithm^(39)^ to quantitatively measure the cellular frequency of various immune subsets in the baseline bulk transcriptome of a) PCV13 and b) PPSV23 generated in this study, and the publicly available Fluzone dataset^(40)^. To this, we utilized the PCV13 single-cell RNA sequencing data with cellular annotations to develop a signature matrix. We subsequently applied this signature matrix to deconvolve the bulk transcriptomes, utilizing default parameters and a permutation set of 100.

Fluzone RNA-seq data: The effect of influenza vaccination on circulating PBMCs from healthy young individuals was studied using a publicly available bulk RNAseq dataset (n=5)(GSE45735)^(27)^. Normalized expression values (RPM) of the samples at Day 0 (baseline) and Day 7 post vaccination were retrieved from the Gene Expression Omnibus database. Fold difference in the expression of constant heavy chain Ig genes were calculated by subtracting the gene expression values at day 7 post-vaccination (since this is the timepoint where changes are the most significant) from the pre-vaccination baseline values. The mean fold difference of the 5 donors analyzed is visualized in the heatmap in log_2_ scale and were compared against same values from our data (day 10-baseline).

Fluzone response data: We considered the non-frail donors in this cohort and studied the association between CD16^+^ NK cell frequency at baseline and responsiveness to fluzone. The considered microarray data captured the PBMC transcriptomes of older adults (non-frail Fluzone R=5; non- frail Fluzone NR=11) in response to Fluzone (GSE59654)^(40)^. We then retrieved the normalized data, removed probes with detection p-value < 0.05 in more than 20% of the samples and mapped the filtered probes to genes. The preprocessed data was then subjected to immune deconvolution using CIBERSORTx, with the parameters described in the CIBERSORTx analysis section. We then used the Wilcoxon Rank Sum test to compare CD16^+^ NK cell frequency in Fluzone R and NR at baseline.

Fluad response data (GSE211560): We re-analysed the scRNAseq data^(41)^ of fluad responders (R, n=3) and non responders (NR, n=3) to investigate the relationship between the baseline frequency of CD16^+^ NK and response to Fluad using Seurat package in R (v4.0.4). We focused on a subset of cohort comprising of Fluad responders (R, n=3) and non-responders (NR, n=3) and retrieved the baseline samples, which were preprocessed using the filters described in the original study^(41)^. We annotated the subclusters based on known markers (**Fig. s9d**), and computed frequency of different immune cells in the total PBMC. To compare the frequency of CD16^+^ NK cells between Fluad R and NR at baseline, we used the Wilcoxon Rank-sum test.

### Statistical tests

P-value calculation for all box-plots were calculated using two-sided Wilcoxon Rank-sum using *stat_compare_means* function from *ggpubr* package (v0.4.0). For scatter plots, Pearson’s *rho* values were reported, and p-values were calculated using *cor.test* from *stats* package (v4.0.4) at default settings.

---

**Fig s1.**
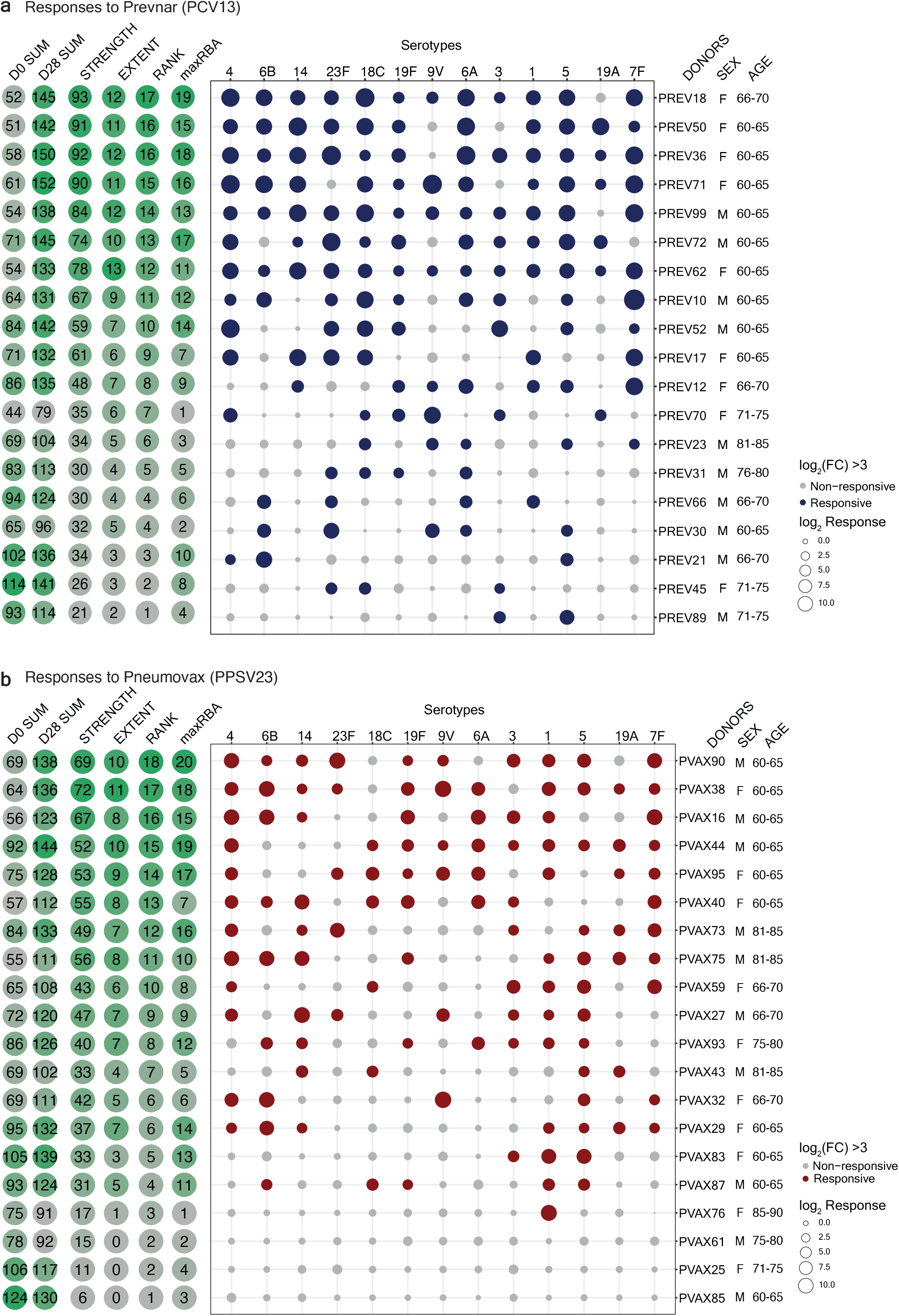
Bubble plot showing the fold change in the antibody titers for individual serotypes in response to PCV13 **a**) and PPSV23 **b**). The size of the dots represent the magnitude of the fold change (FC) value, and the color indicates significant response (Log_2_ FC is > 3): blue for PCV13 and red for PPSV23. Donors are ordered from top to bottom according to the vaccine response Rank. On the left side, D0 SUM (sum of pre-vaccination OPA titers to 13 serotypes), D35 SUM (sum of 28 days post-vaccination OPA titers to 13 serotypes), the Strength, the Extent, the Rank and the maxRBA rank (sum of baseline adjusted fold changes to 13 serotypes) for each donor is displayed.

**Fig s2.**
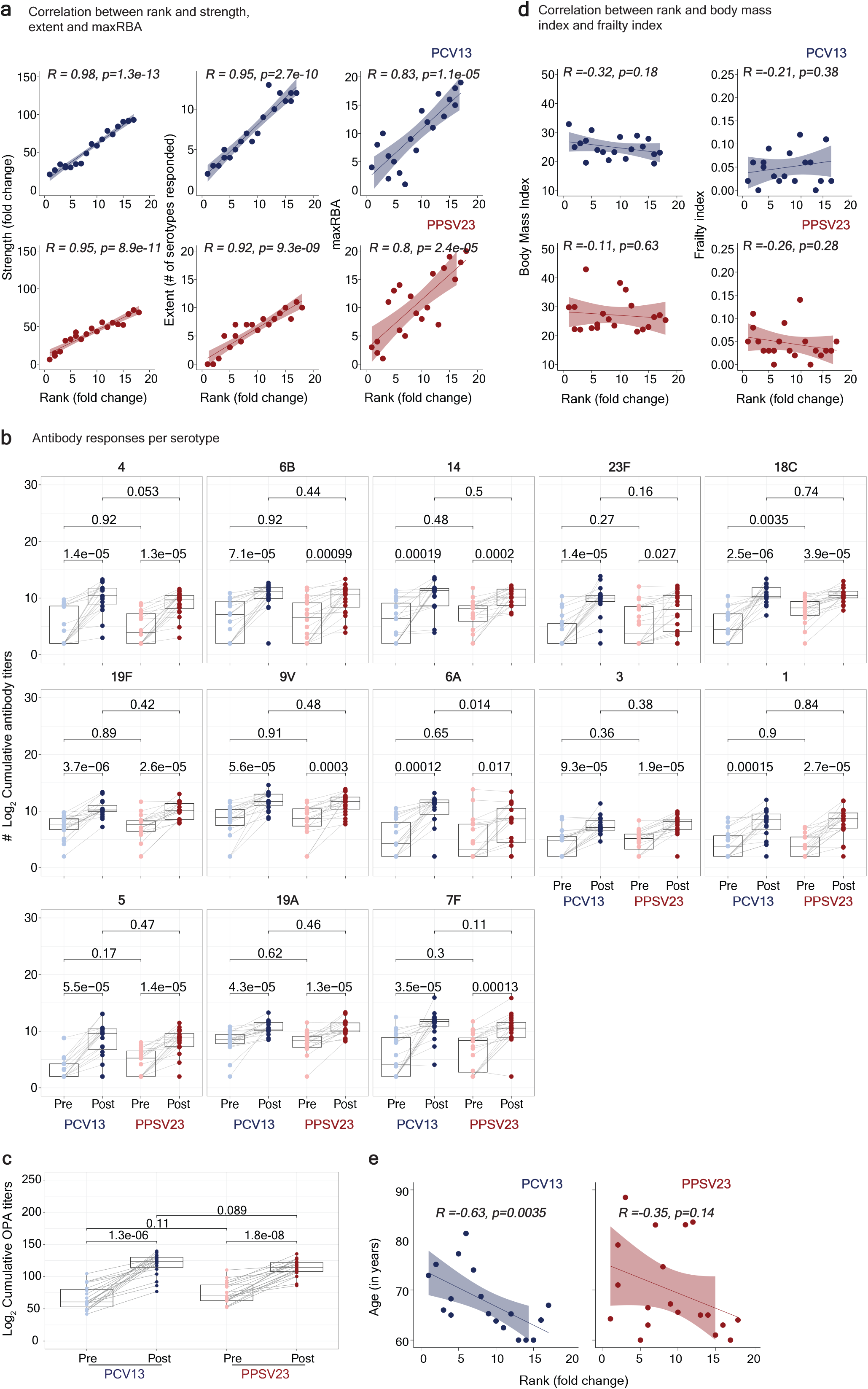
**a**) Correlation between the Rank and the Strength, the Extent, the maxRBA for PCV13 (top panel) and PPSV23 (bottom panel). **b**) OPA titers (Log_2_) for individual serotypes showing the connecting lines between Pre and post-vaccination. **c)** Pre-vaccination and post-vaccination cumulative OPA titers (expressed as sum Log_2_) excluding serotype 6A. **d**) Correlation between the Rank and body mass index (BMI) and frailty index (FI). **e)** Correlation between Rank and age (in years) for PCV13 and PPSV23. The Pearson correlation metric was used to perform correlation analysis (a, c, and d). The Wilcoxon matched-pairs signed-rank test was used in to compare the difference in mean between pre- and post-vaccination OPA titers for 13 serotypes (b) and cumulatively (c) in PCV13 and PPSV23.

**Fig s3.**
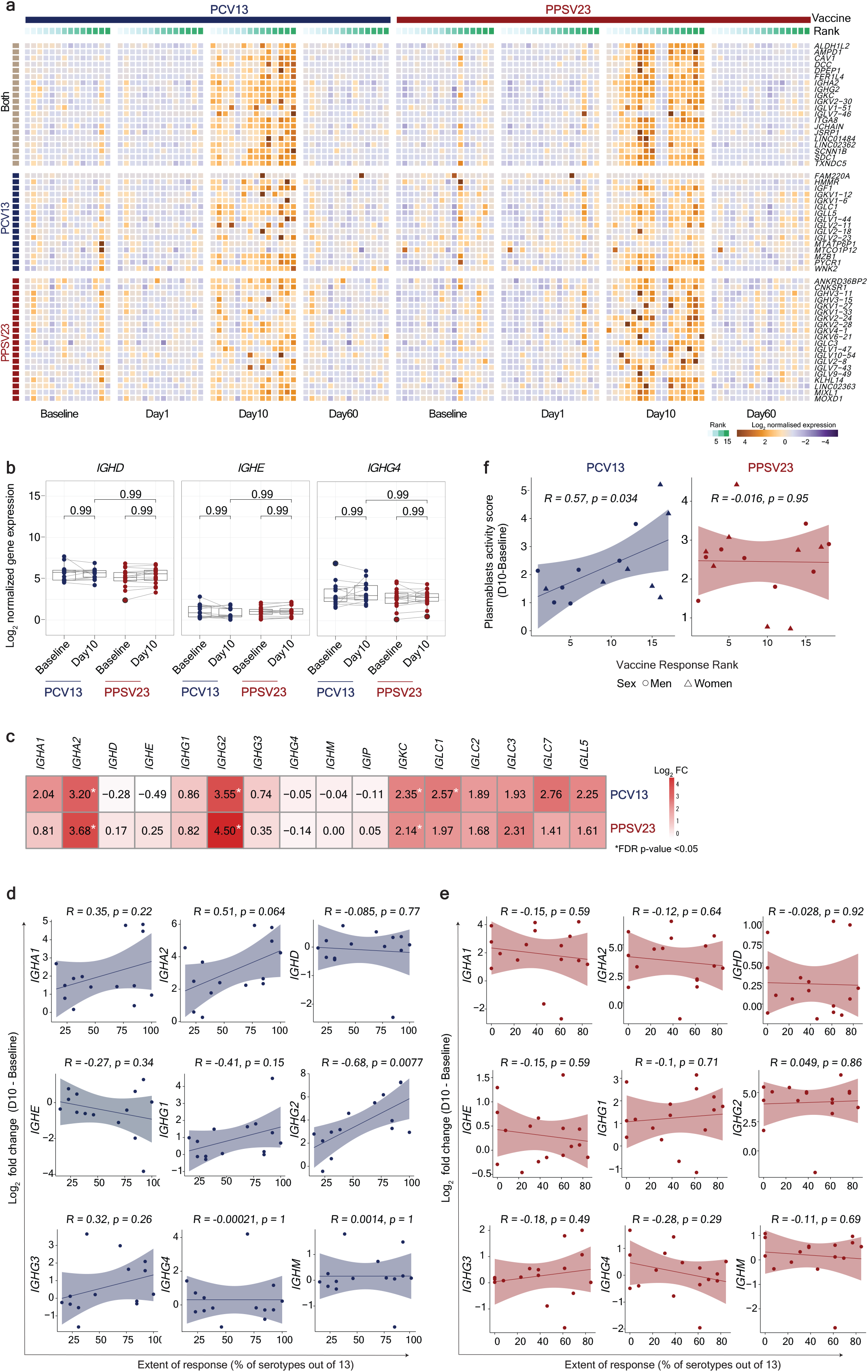
**a**) Heatmap showing differentially expressed genes between Day 10 and baseline, using the normalized gene expression. Differentially expressed genes that are common for both vaccines or show differential expression only in PCV13 or PPSV23 are grouped separately. **b**) Boxplots of the normalized expression of genes coding for the constant region of immunoglobulin heavy chain structure. **c**) Heatmap of differential expressed genes coding for the constant region of immunoglobulin heavy chain structure at Day 10 in response to PCV13 and PPSV23. Genes with a FC >1.5-fold difference and a FDR p-value <0.05 are marked with stars. **d)** Correlation analysis between the fold difference in immunoglobulin genes (gene expression at Day 10 - baseline) and the extent of the response (% of serotypes out of 13) for PCV13. **e)** Correlation analysis between the fold difference in immunoglobulin genes (gene expression at Day 10 - baseline) and the extent of the response (% of serotypes out of 13) for PPV23. **f**) Correlation between the plasmablasts activity scores (Day 10 vs. baseline) and vaccine responsiveness Rank. The Pearson correlation metric was used to perform correlation analysis (a, c, and d). The Wilcoxon matched-pairs signed-rank test was used to compare the expression of immunoglobulin genes at baseline and Day 10 for PCV13 and PPSV23 (c).

**Fig s4.**
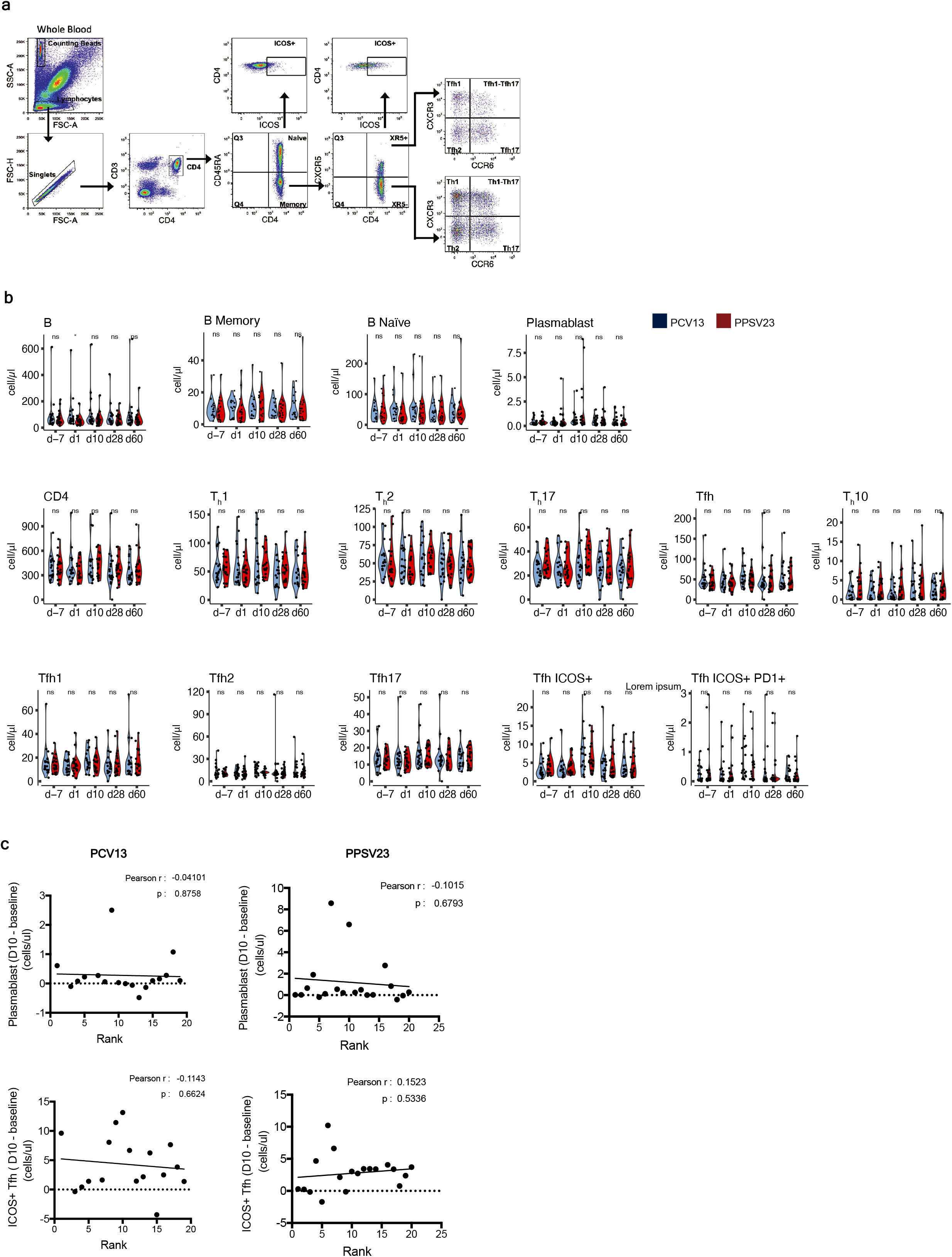
**a**) Representative plots showing the gating strategy for the characterization by flow cytometry of the different memory CD4+ T cell populations. **b**) Comparative analysis of the absolute number of the different cell types between PCV13 and PPSV23 cohorts over time. **c**) Correlation between fold difference in plasmablasts cell numbers (d10 - baseline) and vaccine responsiveness Rank (top panel). Correlation between fold difference in ICOS+ Tfh cell numbers (d10 - baseline) and vaccine responsiveness Rank (bottom panel). The Wilcoxon Rank sum test was used to compare the absolute numbers of different cell types: * (p<0.05), ** (p<0.01), ***(p<0.001), **** (p<0.0001) (b). The Pearson correlation metric was used to perform correlation analysis (c).

**Fig s5.**
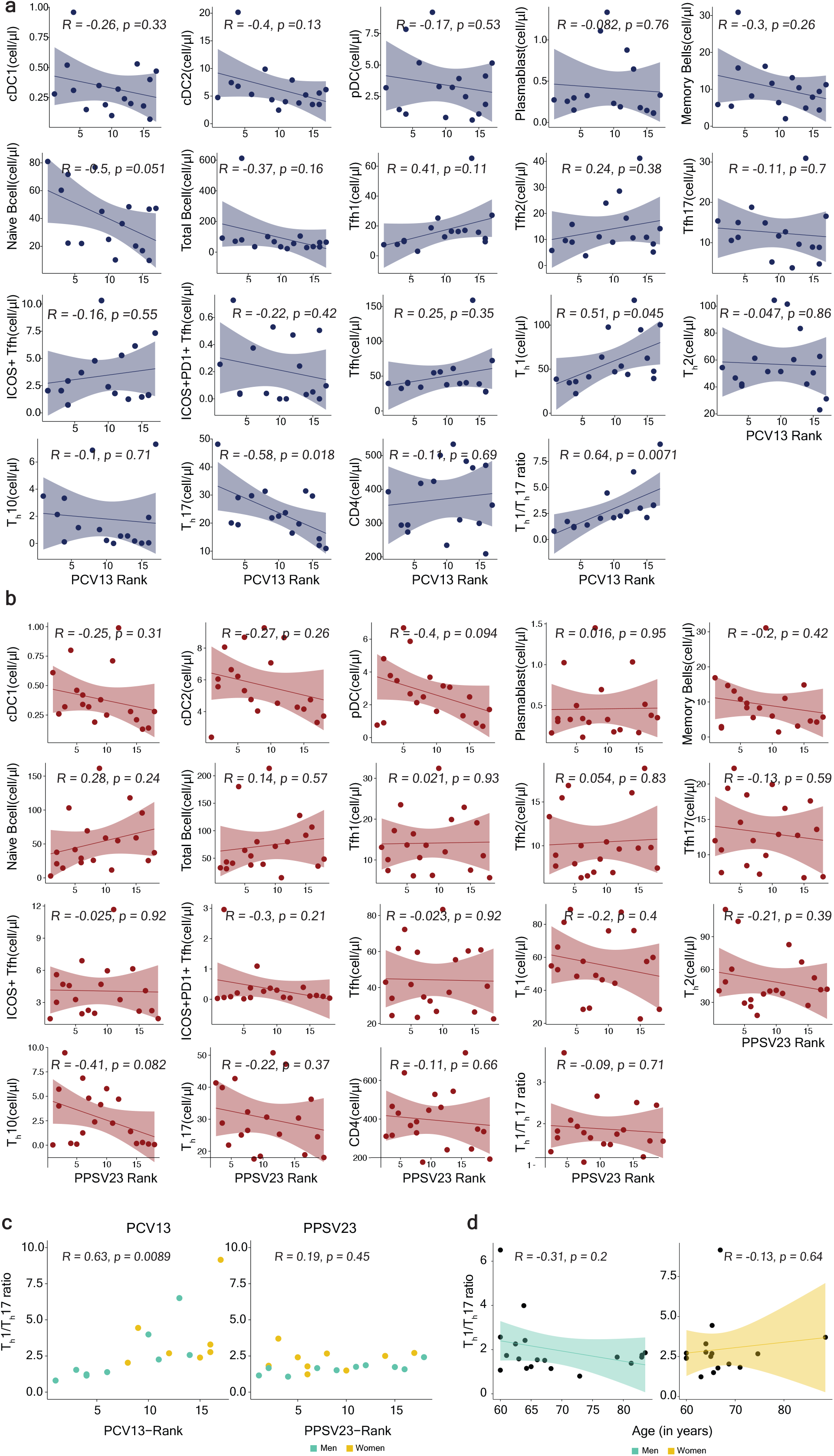
**a**) Correlation analysis between PCV13 Rank and the absolute numbers (cells/ul) of the different cell populations at baseline (d-7). **b**) Correlation analysis between PPSV23 Rank and the absolute numbers (cells/ul) of the different cell populations at baseline (d-7). **c)** Correlation analysis of T_h_1/T_h_17 ratio and vaccine responsiveness Ranks of PCV13 and PPSV23, respectively. **d**) Association between T_h_1/T_h_17 ratio and age among men (green) and women (dark yellow) in response to PCV13 and PPSV23. The Pearson correlation metric was used to perform correlation analysis (a, b, c, and d).

**Fig s6.**
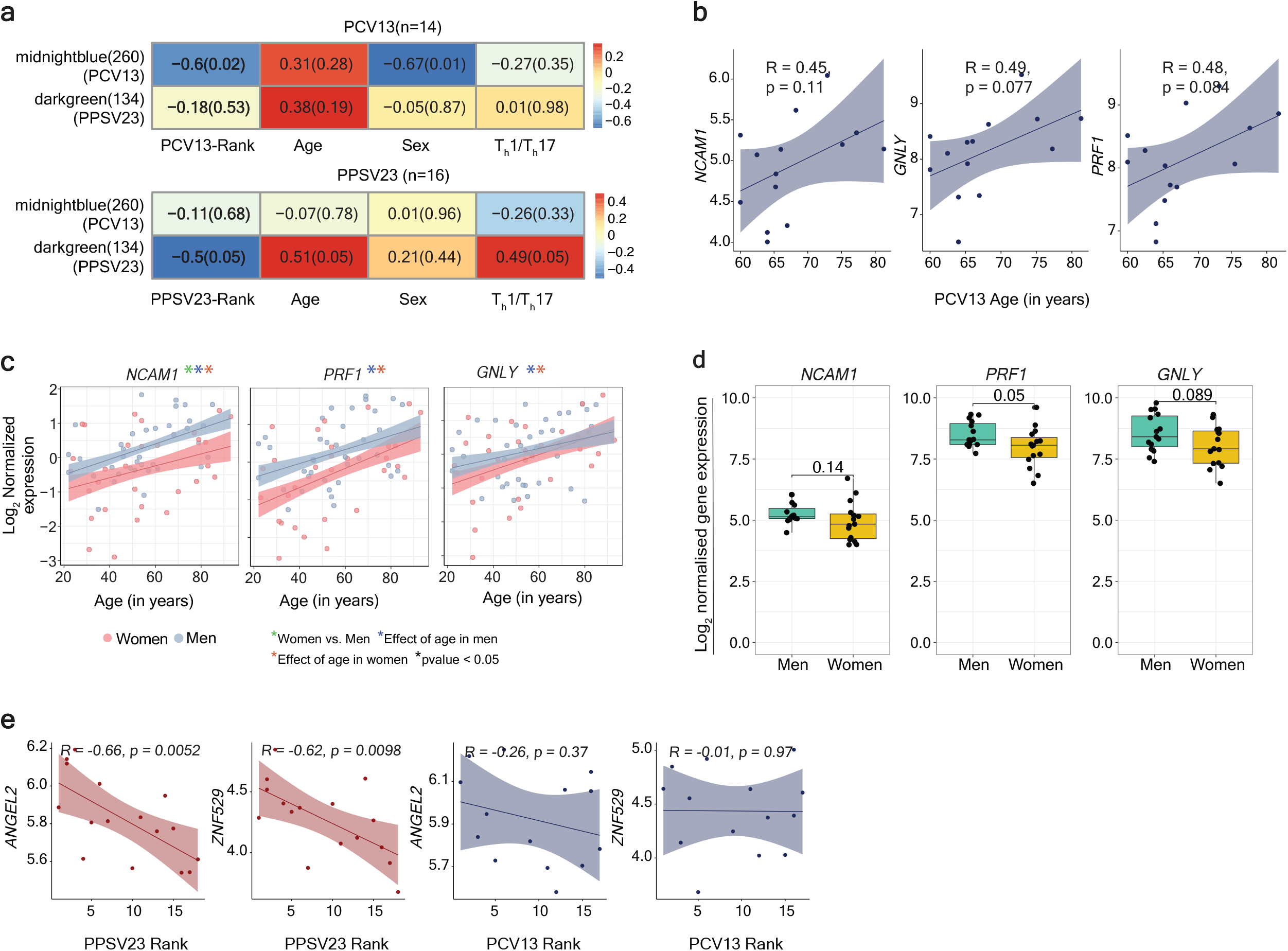
**a**) Heatmap showing the modules correlated with PCV13 Rank, PPSV23 Rank, sex, age and T_h_1/T_h_17 ratio. **b)** Correlation between the expression of cytotoxic genes at baseline and age (in years), in donors who received PCV13 (n=14). **c**) Correlation between the expression of cytotoxic genes and age (in years) in an independent dataset. **d)** Sex differences in the expression of *NCAM1*, *PRF1* and *GNLY* at baseline in both cohorts (n=30). **e**) Correlation analysis between top correlates of darkgreen module (*ANGEL2*, and *ZNF529)* and PPSV23 Rank. Correlation analysis between top correlates of darkgreen module (*ANGEL2,* and *ZNF529)* and PCV13 Rank. Correlation analysis was performed using the Pearson correlation metric.

**Fig s7.**
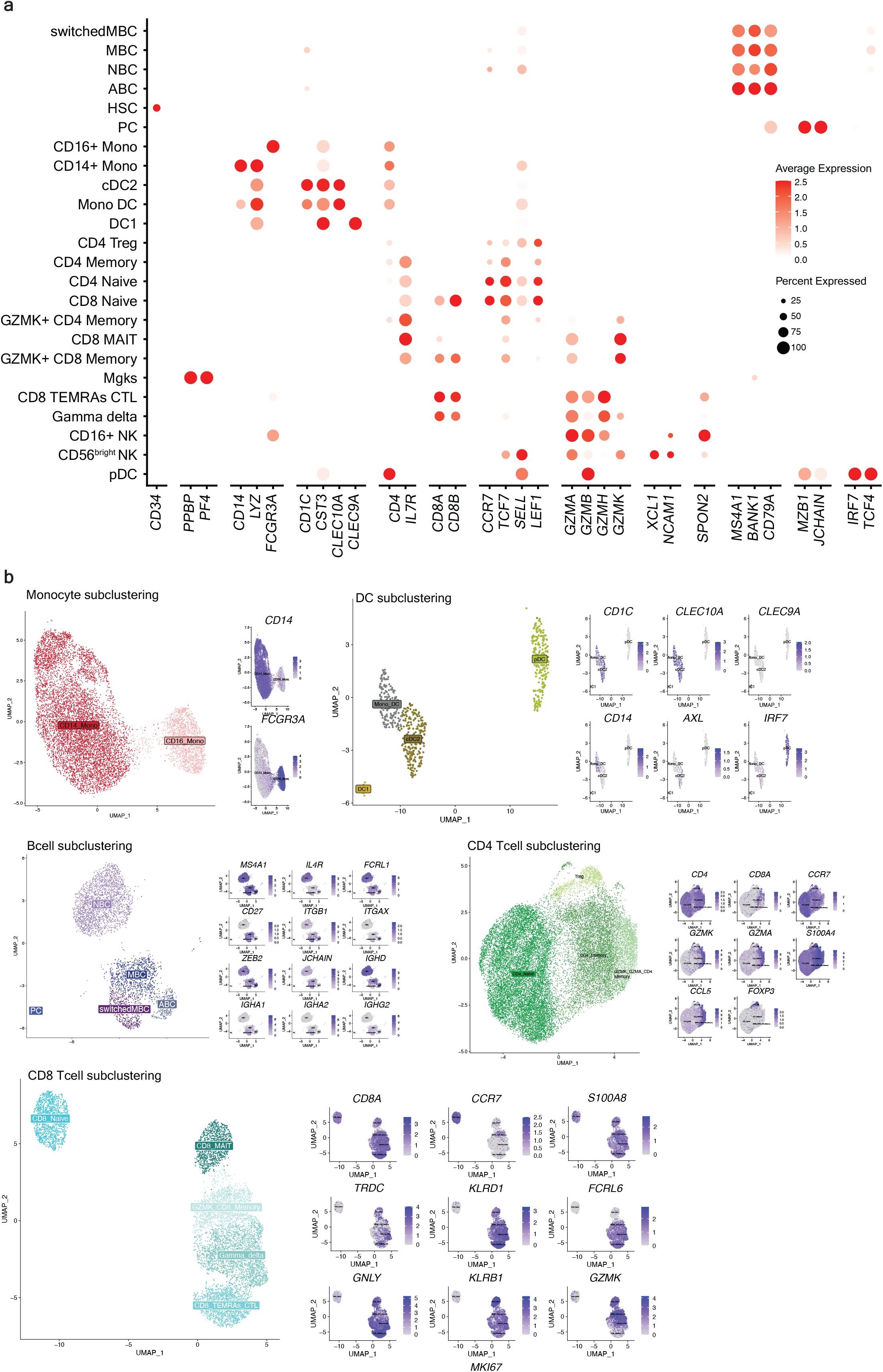
**a)** Dot plot showing the marker gene expression for each subset detected in the PBMC scRNA-seq data**. b)** Single cell subclustering of Monocytes, DCs, B, CD4 and CD8 T cells. Feature plots showing the known markers in blue.

**Fig s8.**
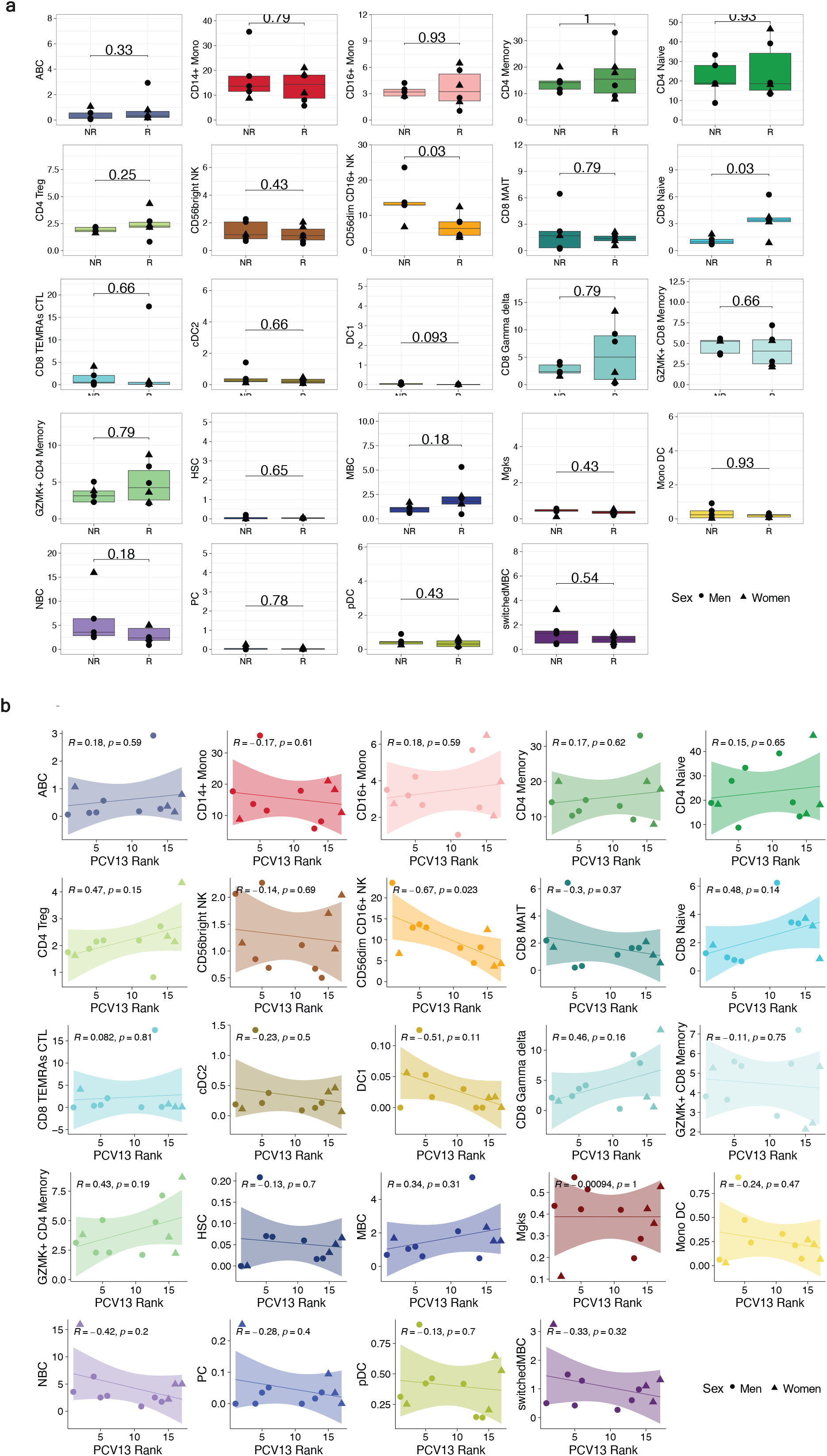
**a**) Stacked bar plot showing immune cell compositions in responders (R) and non-responders (NR). Cell types showing significant difference between R and NR are marked with a star (pvalue < 0.05 starred in red). Boxplot showing the frequency of different immune cell subsets in R and NR. **b**) Correlation analysis between PCV13 rank and the frequency of different immune cell subsets. The Wilcoxon Rank sum test was used to compare the cell percentages between PCV13-R and NR. Correlation analysis was performed using the Pearson correlation metric.

**Fig s9.**
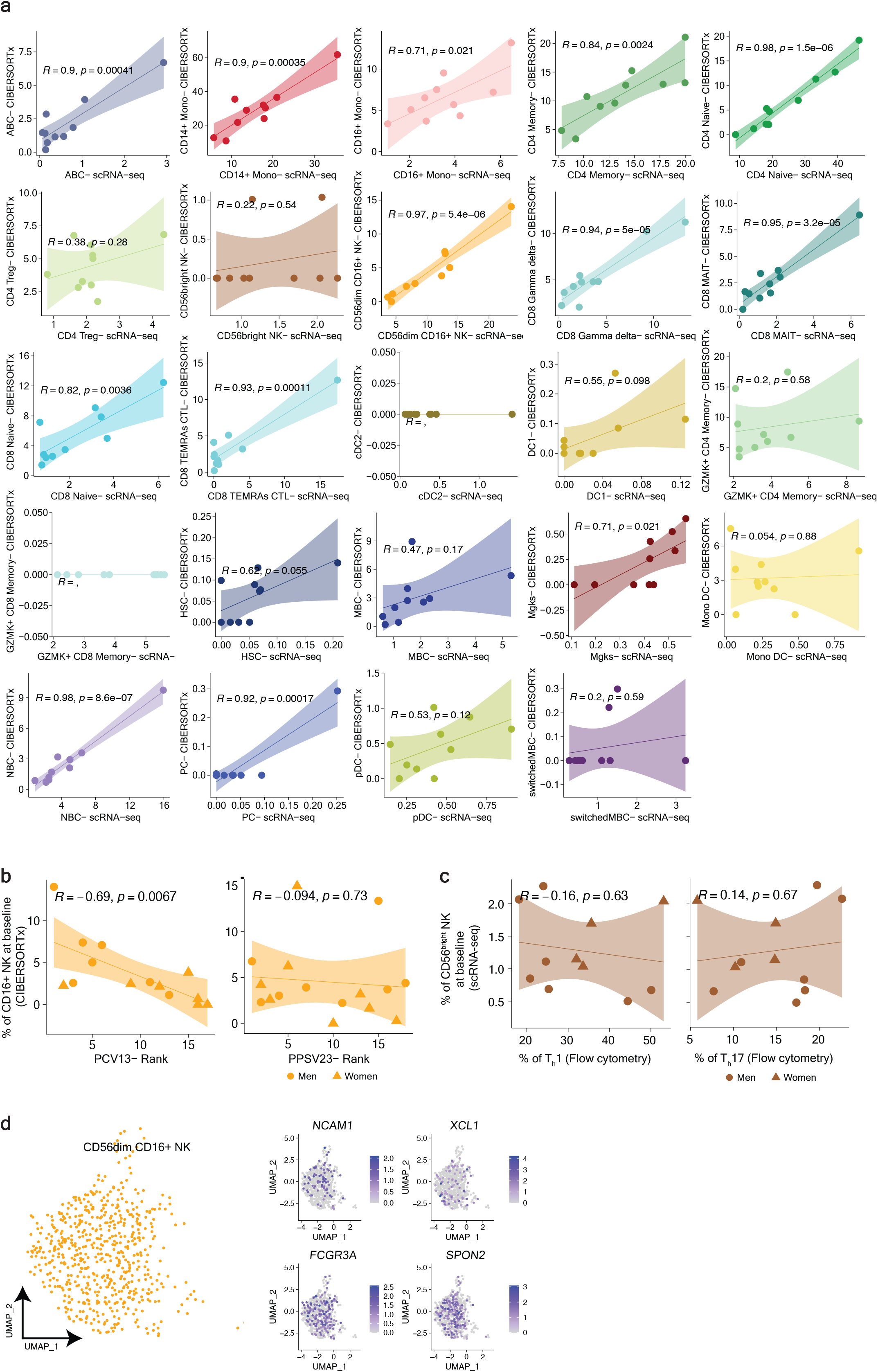
**a**) Correlation between pre-vaccination cell frequency estimated by scRNA-seq and CIBERSORTx for different immune subtypes. **b**) Correlation analysis of CD16^+^ NK frequency at baseline and vaccine responsiveness Ranks of PCV13 and PPSV23, respectively. For the correlation analysis with PPSV23-Rank, cell frequency estimated by CIBERSORTx from the bulk PPSV23 baseline transcriptomes (n=16) was considered. **c**) Correlation analysis of CD56^bright^ NK frequency (n=11) and T_h_1, and T_h_17 frequency at baseline. Pearson correlation was used to perform correlation analysis (a,b,c). **d**) UMAP representation of CD16^+^ NK subsets in fluad responders and non- responders at baseline. Feature plots showing *NCAM1*, *XCL1*, *FCG3RA* and *SPON2* in CD16^+^ NK cells in blue.

